# Neuroimaging correlates and predictors of response to repeated-dose intravenous ketamine in PTSD: preliminary evidence

**DOI:** 10.1101/2021.04.10.21255127

**Authors:** Agnes Norbury, Sarah B. Rutter, Abigail B. Collins, Sara Costi, Manish K. Jha, Sarah R. Horn, Marin Kautz, Morgan Corniquel, Katherine A. Collins, Andrew M. Glasgow, Jess Brallier, Lisa M. Shin, Dennis S. Charney, James W. Murrough, Adriana Feder

## Abstract

Promising initial data indicate that the glutamate N-methyl-D-aspartate (NMDA) receptor antagonist ketamine may be beneficial in post-traumatic stress disorder (PTSD). Here, we explore the neural correlates of ketamine-related changes in PTSD symptoms, using a rich battery of functional imaging data (two emotion-processing tasks and one task-free scan), collected from a subset of participants of a randomized clinical trial of repeated-dose intravenous ketamine *vs* midazolam (total *N*=21). In a pre-registered analysis, we tested whether changes in an *a priori* set of imaging measures from a target neural circuit were predictive of improvement in PTSD symptoms, using leave-one-out cross-validated elastic-net regression models (regions of interest in the target circuit consisted of the dorsal and rostral anterior cingulate cortex, ventromedial prefrontal cortex, anterior hippocampus, anterior insula, and amygdala). Improvements in PTSD severity were associated with increased functional connectivity between the ventromedial prefrontal cortex (vmPFC) and amygdala during emotional face-viewing (change score retained in model with minimum predictive error in left-out subjects, standardized regression coefficient [β]=2.90). This effect was stronger in participants who received ketamine compared to midazolam (interaction β=0.86), and persisted following inclusion of concomitant change in depressive symptoms in the analysis model (β=0.69). Improvement following ketamine was also predicted by decreased dorsal anterior cingulate activity during emotional conflict regulation, and increased task-free connectivity between the vmPFC and anterior insula (βs=-2.82, 0.60). Exploratory follow-up analysis via dynamic causal modelling revealed that whilst improvement in PTSD symptoms following either drug was associated with decreased excitatory modulation of amygdala→vmPFC connectivity during emotional face-viewing, increased top-down inhibition of the amygdala by the vmPFC was only observed in participants who improved under ketamine. Individuals with low prefrontal inhibition of amygdala responses to faces at baseline also showed greater improvements following ketamine treatment. These preliminary findings suggest that, specifically under ketamine, improvements in PTSD symptoms are accompanied by normalization of hypofrontal control over amygdala responses to social signals of threat.

## Introduction

Emerging evidence suggests that intravenous administration of ketamine may also improve symptoms of PTSD – over and above effects on comorbid depression [1,2]. Although the mechanism by which ketamine alleviates mood and stress-related psychopathology is not fully understood, it is thought that this may include promoting synaptogenesis in the prefrontal cortex and hippocampus: thereby reversing characteristic structural deficits observed in these brain regions following chronic stress [3–7]. Further, it has recently been proposed that ketamine administration results in a window of increased neuroplasticity that may support the un- or re-learning of maladaptive associations that contribute to longer-term symptom maintenance in these disorders – for example via facilitating the extinction of fear memories [8,9]. Previous studies in individuals with treatment-resistant depression suggest that improvements in depressive symptom severity following ketamine are associated with changes in neural connectivity during emotional processing (e.g., [10–12]). Dysfunction in a partially overlapping set of brain regions in response to both trauma cues and trauma-unrelated emotional stimuli has previously been identified in PTSD [13–17]: however to our knowledge no previous studies have reported neural correlates of symptom change during ketamine treatment for PTSD.

Here, we analyse a battery of neuroimaging data collected during a recent randomized clinical trial of repeated-dose ketamine *vs* midazolam for chronic, severe PTSD [2]. *N*=21 trial participants consented to provide imaging data before and after treatment, allowing us to preliminarily identify changes in neural functioning that accompany symptom improvement. Given the limited available sample size, we harnessed knowledge derived from previous functional imaging studies to define a set of candidate measures that may *a priori* be expected to be related to both experience of PTSD symptoms and ketamine-related changes in neural function. In addition to this target-driven approach, we used a statistical method appropriate for such datasets (i.e., where the number of observations is small relative to the number of measures) to identify features in this circuit most robustly related to symptom improvement, via a pre-registered analysis. Finally, exploratory follow-up analyses were used to probe symptom specificity and directionality of the target circuit feature most reliably identified in the pre-registered analysis models.

Although this is an observational analysis, meaning we are unable to disambiguate whether changes in brain function following treatment are causally related to or downstream effects of changes in symptom levels, data presented here may help generate testable hypotheses regarding the mechanism of action of ketamine in PTSD when further data becomes available. Specifically, we suggest that improvement in PTSD symptom severity following ketamine is accompanied by normalization of hypofrontal control over amygdala responses to social signals of threat – brain circuitry which has previously been implicated in emotional regulation and extinction learning. If confirmed in future studies, this finding has implications for the potential utility of combining plasticity-promoting agents such as ketamine with psychological therapies that directly encourage reconsolidation and extinction of trauma memories [8,9,18].

## Materials and Methods

### Pre-registration of analysis

A pre-registered analysis plan was deposited with the Open Science Framework (OSF) [19,20]. The point of pre-registration was approximately 2/3 of the way through imaging data collection: at this time both imaging and clinical data were unseen by the primary analyst [20]. Time-stamped pre-registration documents and analytic code used to produce all results reported here are available at the project OSF page (https://osf.io/8bewv/). Follow-up analyses not detailed in the pre-registered analytic plan are described below (see **Exploratory follow-up analyses**).

### Participants

Participants were individuals taking part in a randomized clinical trial of repeated-dose ketamine for chronic PTSD (NCT02397889), results of which are reported elsewhere [2]. All participants met DSM-5 criteria for PTSD and were recruited from the wider trial cohort on an *ad hoc* basis, based on their consent to take part in the scanning sessions, and MRI eligibility criteria (for full inclusion/exclusion criteria see Supplementary Material). All participants gave written consent, and the study received ethical approval from the Institutional Review Board at the Icahn School of Medicine at Mount Sinai.

### Procedures

Study procedures are outlined in **Figure 1a**. Trial participants completed an initial screening visit, then received six intravenous infusions of either ketamine (0.5mg/kg) or the psychoactive placebo control drug midazolam (0.045mg/kg; both three infusions per week for two weeks). Participants and research staff were blind to drug conditions. Imaging data were collected during a baseline (pre-infusion) scan session and a second post-infusion scan following at least one week of treatment (for 2/3 participants this was the day after the 4^th^ drug or 5^th^ infusion, and for 1/3 participants, this occurred within 48 hours of the 6^th^ infusion, due to scheduling conflicts).

**Figure 1.**
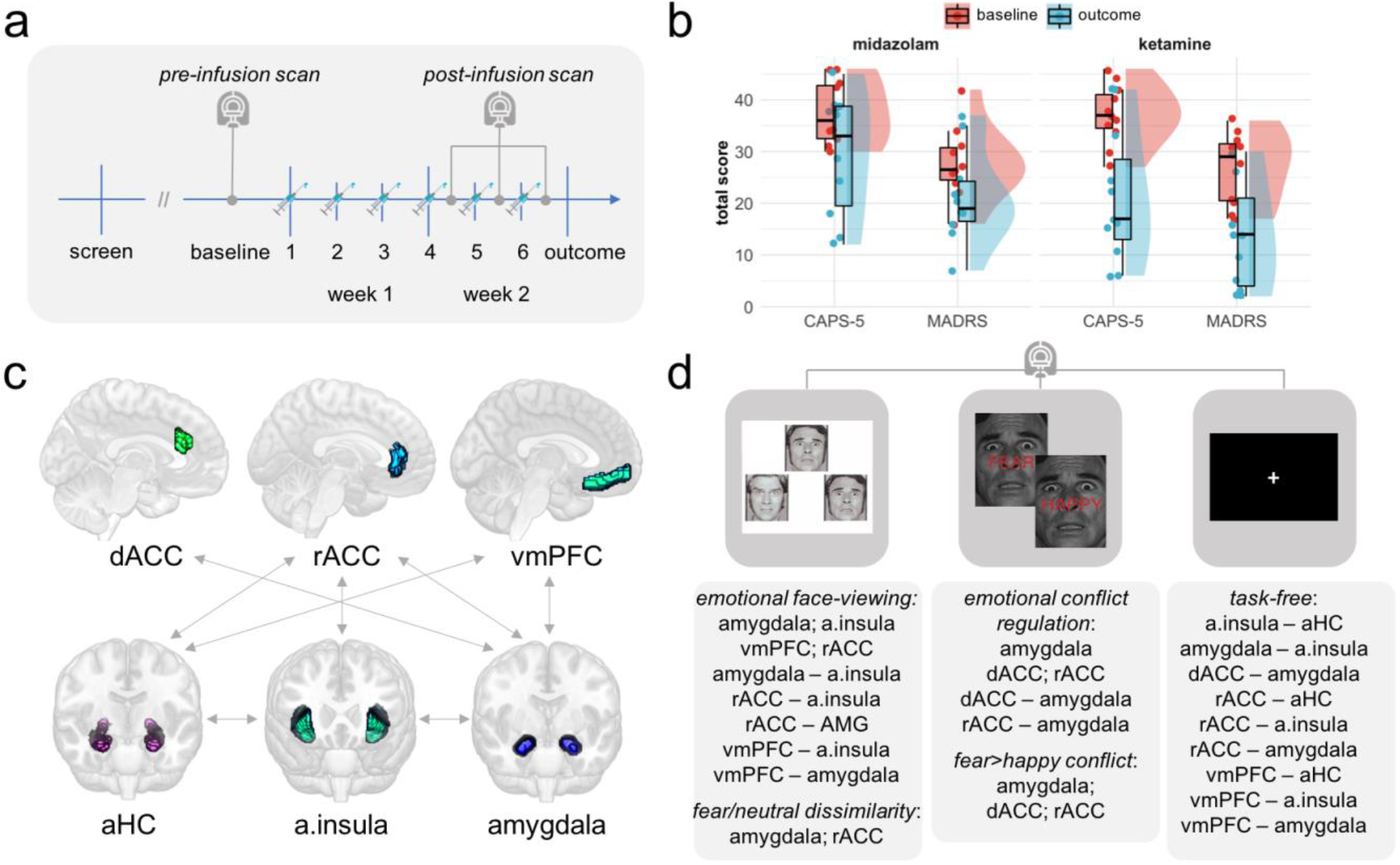
Description of study procedures, clinical measures, and functional imaging measures. **a** Timeline of study procedures. Upon recruitment to the trial, participants were randomly allocated to a drug condition (ketamine vs midazolam) by the trial pharmacist in a 1:1 ratio. Participants received three intravenous infusions per week for two weeks of either 0.045mg/kg midazolam or 0.5mg/kg ketamine. The baseline (pre-infusion) imaging session took place prior to administration of any drugs. The second (post-infusion) scan was the day after the 4^th^ or 5^th^ drug infusion for 2/3 participants, and within 48 hours of the 6^th^ infusion for 1/3 participants. **b** Baseline (pre-infusion 1) and outcome visit clinical measures for all study participants. CAPS-5, Clinician-Administered PTSD Scale for DSM-5; MADRS, Montgomery-Åsberg Depression Rating Scale. Both measures probed symptom levels over the past week and were administered by blinded raters, who were not present during drug infusion sessions. Raincloud plots were generated using [92]. **c** Depiction of the target circuit (regions of interests [ROIs; coloured overlays], and connections between ROIs [grey arrows]) from which functional imaging measures were extracted. dACC, dorsal anterior cingulate cortex; rACC, rostral anterior cingulate cortex, vmPFC, ventromedial prefrontal cortex; aHC, anterior hippocampus; a.insula, anterior insula. **d** Top panels, the three functional imaging measures collected during each scan session: 1) the emotional face-processing task; 2) the emotional conflict regulation (face Stroop) task; 3) 12-minute task-free (resting state) scan. Bottom panels, imaging measures extracted from each functional run. Single ROIs represent mean univariate BOLD signal, pairs of ROIs represent generalized psychophysiological interaction (gPPI) functional connectivity estimates, and the multivariate dissimilarity metric was cross-validated linear discriminant contrast values.

### Clinical measures

The primary clinical measure was the Clinician-Administered PTSD Scale for DSM-5 (CAPS-5; [12]), a structured clinical interview that yields a measure of total symptom severity over the past week (a composite metric of frequency and intensity for each DSM-5 PTSD symptom). Given substantial comorbidity between PTSD and depression, the Montgomery-Åsberg Depression Rating Scale (MADRS; [22]) was also administered. Both measures were administered by blinded raters, who were absent during drug infusion sessions (and therefore unexposed to differences in subjective reaction to study drugs between participants). Information about psychotropic medication status and current self-medication with marijuana or cannabis derivatives was also recorded (see Supplementary Material).

### Drug side-effect measures

Clinician-rated dissociative, psychotomimetic, and manic effects [23–25] and patient-rated somatic effects [26] were included in the analysis of symptom change data, in order to allow for potential effects of functional unblinding of ketamine *vs* midazolam, or non-specific effects related to the magnitude of psychoactive reaction to either drug, in symptom change data [27,28] (see Supplementary Material).

### Neuroimaging data

On each imaging session, a T1-weighted structural and three T2*-weighted functional imaging runs were collected. Functional runs consisted of: 1) an emotional face-processing task [29]; 2) an emotional conflict regulation (‘face Stroop’) task [30]; and 3) a 12-minute eyes-open task-free (‘resting state’) scan (**Figure 1c**).

#### Task descriptions

The emotional face-processing task was a paradigm frequently used in imaging studies of emotional processing [29]. On each trial, participants view a triplet of either emotional faces or ovoid shapes, and are required to make a simple perceptual judgement (which of a left-right pair matches the stimulus in the center of the screen). Trials were ordered according to a block design with interleaving blocks of shapes trials and emotional faces trials (six trials per block, one block each of neutral, fearful, surprised and angry faces). The emotional conflict regulation (face Stroop) paradigm was chosen on the basis of previous sensitivity to PTSD status [30]. On each trial, participants view an emotion word (“happy” or “afraid”), superimposed on a photograph of an emotional face (happy or afraid). Participants must respond according to the emotion displayed by the face, ignoring the meaning of the superimposed word. On congruent (low-interference) trials, the background face displays the same emotion as the superimposed word, and on incongruent (high-interference) trials, the target face displays the alternative emotion. Stimuli were presented in pseudorandom order with jittered presentation timing, in an event-related design (60 congruent, 60 incongruent, and 60 baseline trials). During the task-free scan, participants viewed a fixation cross on the screen, and were asked to keep their eyes open.

#### Data acquisition

Imaging data were acquired using a 3T Siemens Biograph mMR scanner (Siemens Healthcare) with 64-channel head coil. T2*-weighted images were collected using an echoplanar imaging (EPI) sequence (TR=2000ms, TE=0.027ms, flip angle=90°, slices per volume=38, voxel size 2×2×2mm). The T1-weighted structural scan used an MPRAGE sequence (voxel size 1×1×1mm).

#### Pre-processing

Imaging data were pre-processed using fMRIprep, version 1.1.4 [31,32]. Briefly, images were slice-time corrected, spatially normalized, and smoothed with an isotropic Gaussian kernel of 6mm FWHM, followed by non-aggressive independent components analysis (ICA-AROMA) denoising [33] and re-sampling to standard MNI space (for full details see Supplementary Material).

#### First level analysis

First level models were specified using SPM12 (Wellcome Trust Centre for Neuroimaging, 2014), run in MATLAB, version R2019a. For the emotional face-processing task, onsets of emotional faces and comparison shape-judgement trials were modelled separately. For the emotional conflict regulation task, onsets of incongruent and congruent trials were modelled separately. Primary contrasts of interest were faces>shapes and incongruent>congruent trials, respectively. For both tasks, erroneous responses were included as regressors of no interest in the calculation of these contrasts, with onsets modelled at the time of response. Nuisance regressors describing mean time-series and spatially-coherent noise components in white matter and cerebrospinal fluid [34], plus outlier volumes based on movement and intensity traces [35] were also included in all first level models (for full specification see Supplementary Material).

#### Target circuit definition

Due to our relatively small sample size, we decided to restrict our analysis to measures drawn from an *a priori* target circuit, defined on the basis of previous task-based functional imaging studies in individuals with PTSD ([13–16]; see [17] for a recent meta-analysis). This target network consisted of the ventromedial prefrontal cortex (vmPFC), rostral and dorsal anterior cingulate cortices (rACC and dACC), anterior insula, amygdala, and anterior hippocampus (aHC) (**Figure 1d**). Notably, signal changes in or between several of these brain regions during emotional processing tasks have been also identified following intravenous ketamine in healthy volunteers and individuals with treatment-resistant depression [10–12,36].

#### Mask definition

Masks for anatomically well-defined regions of interest (ROIs) (amygdala, anterior insula) were generated using the Neuromorphometrics maximum probability tissue label atlas distributed with SPM12. Masks images for other ROIs (vmPFC, rACC, dACC, aHC) were generated from meta-analytic consensus maps using Neurosynth [37] (see Supplementary Material).

#### Derivation of imaging measures

For each functional scan, a pre-defined set of imaging measures (mean regional BOLD signal, covariation in time series or functional connectivity between regions, and/or multivariate representational similarity across voxels within a region) were extracted. Briefly, mean BOLD signal estimates across ROIs were extracted from the contrast of interest for each first level model using MarsBaR [38]; task-specific ROI-ROI covariances or functional connectivity estimates were generated via generalized psychophysiological interaction (gPPI) analysis using the CONN toolbox [39]; and multivariate representational similarity metrics (linear discriminant contrast estimates) were generated using the RSA toolbox [40], run in MATLAB (see Supplementary Material). The full set of imaging measures submitted to further analysis is listed in **Figure 1c**.

In order to further limit chance of Type 1 error related to across- or within-subjects differences in motion during scan sessions (which may covary effects of interest, e.g., clinical severity; [31,27]), all imaging measures were first regressed against mean framewise displacement (head movement) recorded during the relevant functional scan. Residuals from these regressions were then passed on to further analysis (*NB*, in the absence of a relationship between within-scan motion and a given imaging metric, the rank ordering of individual estimates should be unaffected by this process; see e.g., [43]). For clarity, all bivariate plots presented in the Results depict ‘raw’ (unregressed/unimputed) values.

### Statistical analysis

Further statistical analysis was carried out in R, version 3.6.1 (R Core Team, 2019). Given our small *N* and lack of previous data regarding the effect of ketamine administration on neural function in PTSD, our primary (pre-registered) analysis was motivated by identifying which of a candidate set of imaging features were more prominently related to PTSD symptom improvement in our dataset. Subsequently, imaging measures and other relevant sociodemographic variables were related to change in PTSD symptom severity following treatment using elastic net regression [44], as implemented in the R package glmnet, version 3.0-2 [45]. Elastic net is a form of penalized regression which balances LASSO (or L1-norm) penalization [46], which forces small effects to zero (yielding feature selection), with ridge (or L2-norm) penalization, which shrinks effects towards the null, and deals better with multicollinearity among (sets of) predictors than LASSO alone [47]. As such, the elastic net is able to cope relatively well with situations where the number of observations is small with respect to the number of measurements, and where measurements are interrelated [44]. Parameters governing balance between L1 and L2-norm penalization (alpha) and degree of regularization applied (lambda) were chosen via grid search and a leave-one-(subject)-out cross-validation (LOOCV) procedure. Specifically, parameter values defining the winning model were chosen so as to minimize error when predicting the target variable in left-out subjects (minimum mean squared error [MSE] model), a process designed to guarding against over-fitting (but see Discussion). Under a prediction framework, retention in the minimum MSE model implies that values of a particular variable may be useful in estimating the target variable (here, change in PTSD symptom severity following treatment) – although there is no inference on significance *per se* [48].

The first pre-registered analysis related *changes* in imaging measures to changes in CAPS-5 total scores following treatment, in order to identify correlates of symptom change. Changes in imaging measures were calculated as post-minus pre-infusion scan metric values, following regression of each metric against mean within-scan movement (see above). The second pre-registered analysis related *baseline values* of imaging measures and other variables to changes in CAPS-5 scores, in order to identify predictors of treatment response. In both cases, a set of nested models was constructed: the first examining at predictors of symptom change across all subjects (drug-agnostic analysis), and the second including interaction terms between imaging measures and a dummy variable encoding received drug identity ([0,1] for midazolam *vs* ketamine), in order to identify effects that were stronger in individuals who received ketamine. A third model also included changes in depressive symptoms (MADRS total scores) following treatment, in order to attempt to identify predictors specific to cardinal PTSD symptomatology (but see below).

All the above models included a set of demographic and clinical measures that may potentially confound imaging or symptom change metrics, specifically: age, gender identity, self-reported education level, concurrent use of stable doses of psychotropic medication, urine toxicology tetrahydrocannabinol (THC) results, four drug side-effects measures (indexing dissociative, psychotomimetic, manic, and somatic effects experienced across infusion sessions), and baseline PTSD severity (CAPS-5 total score). The baseline prediction model also included several other variables which have been previously related to clinical response to ketamine or resilience to trauma-related psychopathology in general, specifically: self-reported income, perceived social support, cognitive test (CogState battery) performance, history of alcohol use disorder in first degree relatives, and dissociative symptoms during the first infusion (see Supplementary Material). As with the imaging variables, the model construction process allowed for retention of any of these additional variables as predictors of symptom change scores, given a parameter weight sufficient to survive penalization, and a reliable contribution to the target outcome across individuals (as determined via LOOCV).

All predictors were scaled (z-scored) prior to entering into the prediction models, yielding standardized regression (β) weights. Symptom change scores were positively framed (calculated as baseline minus outcome visit scores), in order to aid interpretability of model output. Further information regarding sample size, missing data, and deviations from the pre-registered analysis plan is available in the Supplementary Material.

#### Exploratory follow-up analyses

Imaging features identified by the pre-registered analyses as being reliably related to symptom change scores were further investigated using non pre-registered exploratory follow-up analyses, in order to investigate symptom-specificity and directionality of effects in our data.

Specifically, as PTSD is a heterogenous disorder [49,50], exploratory correlation analyses were conducted to examine how strongly changes in imaging metrics were related to changes in different PTSD symptom dimensions, as defined by Armour et al., [49,51]. Given the considerable conceptual overlap between some dimensions (e.g., negative affect, anhedonia) and symptoms of depression, this may be a cleaner way of examining specificity of effects to different aspects of PTSD symptomatology than our pre-registered plan of attempting to remove variance related to change in total depression severity (MADRS) score.

In order to gain insight into the directionality of functional connectivity effects identified in the main analysis, follow-up effective connectivity analysis was carried out via dynamic causal modelling (DCM) [52]. As we lacked sufficient power to specify connectivity structure in a data-driven manner, model structure was informed by previous DCM studies of the same task (in both healthy volunteer and anxiety samples; [53–56]), and limited to bidirectional connectivity between the pair of ROIs specified in the functional connectivity measure. For the relevant pair of ROIs, changes in effective connectivity between pre- and post-infusion imaging sessions that were related to change in PTSD symptoms over the course of treatment, and interactions of these changes with the factor of drug (ketamine vs midazolam), were identified using hierarchical Parametric Empirical Bayes (PEB) analysis [57,58], as per [59] (see Supplementary Material).

## Results

### Study participants and clinical measures

Study participants are described in **Table 1**. Participants had severe, chronic PTSD (mean baseline CAPS-5 score of 37, mean duration 17 years). The majority of participants were women, and the most common primary trauma was sexual violence, followed by other forms of interpersonal violence or abuse. Improvements in PTSD and depressive symptoms over the course of treatment were observed in individuals who received both midazolam (*N*=10) and ketamine (*N*=11); however, greater improvement was observed in the ketamine group: both in the wider clinical trial [2] and the imaging subsample (significant session*drug interaction on CAPS-5 and MADRS total scores in repeated-measures ANOVA with factors of drug, session, and measure; *F*_1,57_=6.58, *p*=0.013; **Figure 1b**; Supplementary Results). In general, response accuracy and timing on the emotion-processing tasks did not vary according to imaging session or received drug, and accuracy was close to ceiling at both pre- and post-infusion imaging sessions (Supplementary Results; **Figure S1**). Distributions of clinical outcome and side-effects variables, and correlation matrices depicting relationships between variables, are available in the Supplementary Material (**Table S1; Figure S2,7**).

**Table 1.** Summary of demographic and clinical variables for study participants, by received drug treatment (ketamine *vs* midazolam). Values represent mean (SD) unless otherwise specified. Low-frequency gender identities and index trauma types were collapsed into other categories, in order to preserve participant privacy. CAPS-5, Clinician-Administered PTSD Scale for DSM-5 (a measure of past-week PTSD symptom severity); MADRS, Montgomery-Åsberg Depression Rating Scale (a measure of past-week depressive symptom severity). *p* values represent the result of statistical comparisons for each variable between drug groups (for continuous variables, this was via Welch’s 2-sample *t* tests; for categorical variables, this was via *χ*^2^ tests).

|  | midazolam<br>(N=10) | ketamine<br>(N=11) | *p* |
| --- | --- | --- | --- |
| Age (years) | 42.3 (12.4) | 42.5 (14.1) | 0.98 |
| Gender (N female, other, or non-binary) | 8 (80%) | 10 (91%) | 0.51 |
| Race (N) |  |  |  |
| Black or African American | 2 (20%) | 1 (9%) | 0.73 |
| Asian | 2 (20%) | 2 (18%) |  |
| White or Caucasian | 4 (40%) | 6 (55%) |  |
| Other | 2 (20%) | 2 (18%) |  |
| Ethnicity (N) |  |  |  |
| Hispanic/Latinx | 3 (30%) | 3 (27%) | 1.0 |
| Education level (N) |  |  |  |
| High school graduate | 0 (0%) | 2 (18%) | 0.56 |
| Some college or trade school | 3 (30%) | 3 (27%) |  |
| Graduated 4-year college | 4 (40%) | 2 (18%) |  |
| >4 years college or professional school | 3 (30%) | 4 (36%) |  |
| Baseline CAPS-5 (past week) total score | 37.6 (6.2) | 37.2 (5.7) | 0.87 |
| Baseline MADRS (past week) total score | 27.8 (7.1) | 27.0 (6.8) | 0.79 |
| PTSD duration (years) | 16.3 (7.2) | 18.5 (19.8) | 0.74 |
| Primary (index) trauma (N) |  |  |  |
| Sexual violence | 4 (40%) | 6 (55%) | 0.44 |
| Physical violence or abuse | 3 (30%) | 3 (27%) |  |
| Witnessed violence or death; combat exposure | 2 (20%) | 2 (18%) |  |
| Medication status (N) |  |  |  |
| Psychotropic medication (stable dose) | 5 (50%) | 6 (55%) | 1.0 |
| Marijuana or cannabis derivative use | 0 (0%) | 3 (27%) | 0.25 |
| Psychological treatment status (N) |  |  |  |
| Currently receiving talk therapy or counselling | 5 (50%) | 5 (45%) | 1.0 |
| Number of days between baseline (pre-infusion) and post-infusion MRI scan | 34.6 (26.3) | 16.1 (7.1) | 0.07 |
| Number of infusions received at time of post-infusion MRI scan | 5.2 (0.8) | 4.7 (0.9) | 0.22 |

### Neuroimaging correlates of PTSD symptom improvement

Across all study participants, the strongest correlate of improvement in PTSD symptom severity retained in the minimum MSE model was increased functional connectivity between the vmPFC and amygdala during emotional face-viewing (standardized regression weight [β]=2.90). Increases in rACC BOLD during negative emotional conflict regulation, and greater task-free (resting) connectivity between the rACC and anterior insula were also retained in the model (βs=0.97, 0.68, respectively; whole model *r*^2^=0.318) (**Figure S3**). When interactions between drug and imaging measures were entered into the model, the interaction term between drug and vmPFC-amygdala connectivity during emotional face-viewing was also retained: indicating a stronger effect in individuals who received ketamine (β=0.86; **Figure 2a,b**). Interactions between drug and dACC BOLD during emotional conflict regulation, and task-free vmPFC-anterior insula connectivity were also retained (βs=-2.82, 0.60; whole model *r*^2^=0.585). Inspection of bivariate correlation plots revealed that improvements in PTSD symptom severity were associated with decreased dACC BOLD during emotional conflict and increased resting vmPFC-anterior insula connectivity, only in individuals who received ketamine (**Figure 2c,d**). In a follow-up analysis which included concurrent change in depressive symptom severity (MADRS total score), the overall effect of increased vmPFC-amygdala connectivity during emotional face-viewing and the interaction of drug with decreased dACC BOLD during emotional conflict regulation were retained in the minimum MSE model (βs=0.69, -0.33; whole model *r*^2^=0.667) – suggesting some specificity of these effects to improvement in cardinal PTSD symptoms (**Figure S4**).

**Figure 2.**
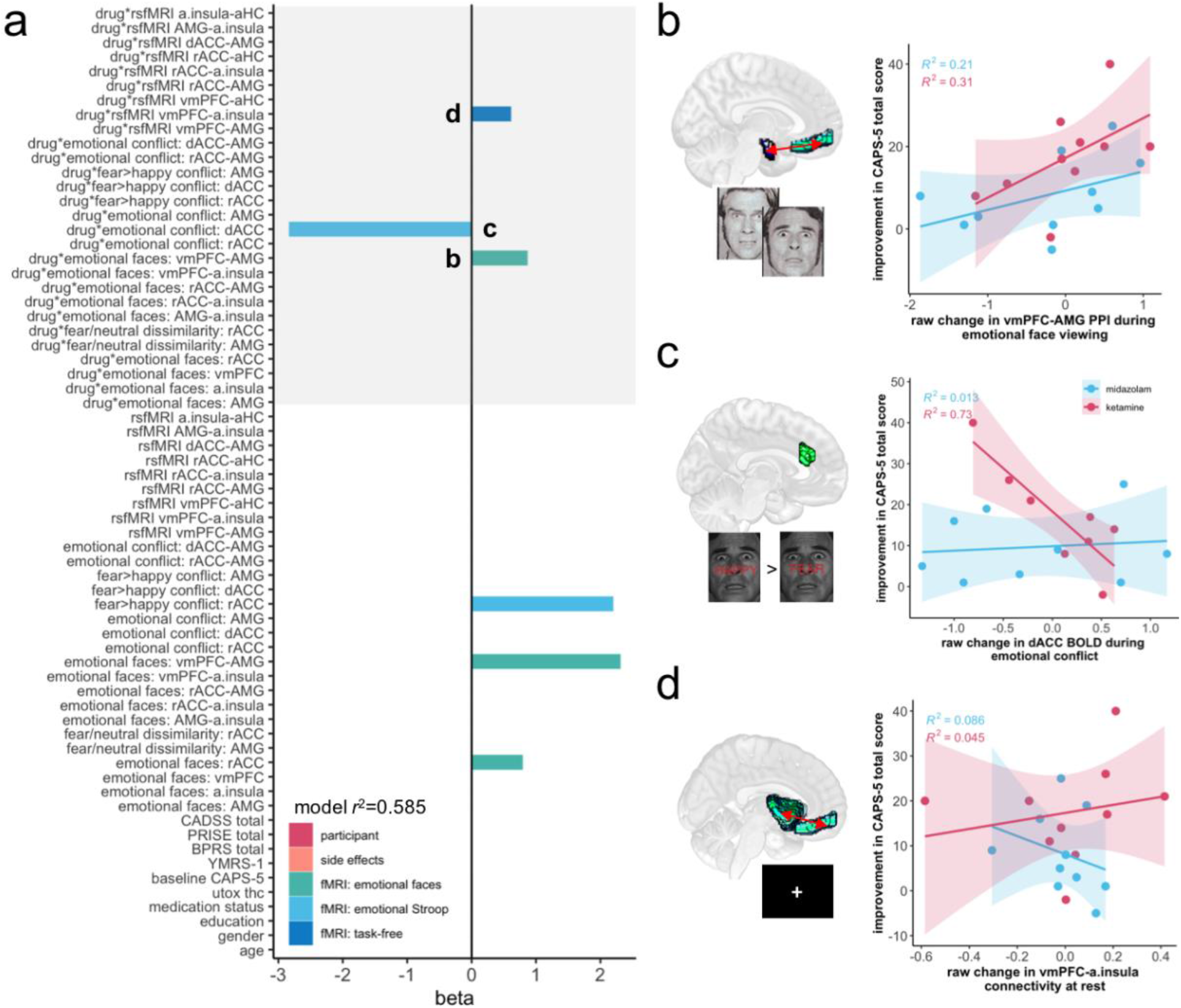
Neuroimaging correlates of PTSD symptom change. **a** Standardized regression coefficient (beta) values for the elastic net model with minimum predictive error for change in PTSD symptoms over the course of treatment in left-out subjects. Non-zero coefficients represent measures with predictive value for change in PTSD severity. All imaging measures represent change scores (post-infusion scan minus pre-infusion scan). Grey shading highlights interaction terms between imaging measures and received drug identity (ketamine vs midazolam). rsfMRI, resting-state/task-free fMRI scan; CADSS, Clinician-Administered Dissociative States Scale; BPRS, four items from the Brief Psychiatric Rating Scale probing psychotomimetic symptoms; YMRS, a single item from the Young Mania Rating Scale indexing elevated mood; PRISE, Patient Rated Inventory of Side-Effects (total score calculated by summing across all somatic domains); CAPS-5, Clinician-Administered PTSD Scale for DSM-5; utox thc, urine toxicology results for presence of THC. **b** Increased connectivity between the vmPFC and amygdala (AMG) during emotional face-viewing was predictive of improvement in total PTSD symptom severity across all participants, but the effect was stronger in individuals who received ketamine. Decreased dACC BOLD during emotional conflict regulation (**c**), and increased resting (task-free) vmPFC-anterior insula functional connectivity (**d**), were only associated with improvement in CAPS-5 total score in individuals who received ketamine. For generalized psychophysiological interaction (gPPI) and task-free functional connectivity estimates, values represent Fisher-transformed correlation coefficients. For regional BOLD signal, values are in arbitrary units. For ease of interpretation, bivariate plots represent ‘raw’ imaging measure values (i.e., calculated prior to regression against average within-scan framewise displacement).

#### Exploratory follow-up analysis by PTSD symptom dimension

An exploratory follow-up analysis was carried out to investigate how changes in the measure most consistently identified across models as a correlate of overall symptom improvement (face-related vmPFC-amygdala connectivity) were related to changes in symptoms across different PTSD symptom dimensions. Increased vmPFC-amygdala coherence during viewing of emotional faces was most strongly related to improvement in re-experiencing, avoidance, and anxious arousal symptoms (*r*s=0.53, 0.55, 0.56, respectively) – with weaker relationships to improvement in negative affect, anhedonia, externalizing behaviour, and dysphoric arousal symptom dimensions (*r*s=0.19–0.45; **Figure S5**). There was no evidence of a relationship between pre-post change in vmPFC-amygdala connectivity during emotional face-viewing and either number of infusions received at the time of the post-infusion scan (Spearman’s ρ=-0.06, *p*>0.5; **Figure S6a**) or number of days between the pre- and post-infusion scan sessions (Spearman’s ρ=-0.29, *p*>0.2; **Figure S6b**).

#### Exploratory follow-up analysis via Dynamic Causal Modelling

Effective connectivity analysis was next used to explore the directionality of vmPFC-amygdala connectivity changes over the course of treatment associated with PTSD symptom improvement. When considering data only from the pre-infusion (baseline) session, PEB analysis of mean effective connectivity across participants revealed strong evidence that the onset of emotional face stimuli modulated the amygdala→vmPFC pathway towards an excitatory connection (posterior estimate [Ep]=0.89, posterior probability [Pp]=1.00), with weaker evidence for face-induced inhibition of the amygdala by the vmPFC (Ep=-0.18, Pp=0.81) (**Figure 3a**; **Table S2**). When considering data from both pre and post-infusion sessions (regardless of drug received), 2^nd^ level PEB analysis revealed that greater improvements in PTSD symptom severity over the course of treatment were associated with shifts towards lesser face-related excitation in the amygdala→vmPFC pathway (Ep=-0.022, Pp=0.98), and greater face-related inhibition in the vmPFC→amygdala pathway (Ep=-0.052, Pp=1.00), post *vs* pre-treatment (**Figure 3b; Table S3**). When separated by drug, there was evidence for an association between greater symptom improvement and lower face-related excitation of the vmPFC by the amygdala in individuals who received both midazolam and ketamine (Ep=-0.019, -0.031, Pp=0.94, 1.00; respectively). However, the relationship between PTSD symptom improvement and greater top-down inhibition of the amygdala by the vmPFC was only evident in the ketamine group (Ep=0.004, -0.091, Pp=0.60, 1.00) (**Figure 3c; Table S3**). Formal comparison between drug conditions via passing the two models to a 3^rd^ level PEB analysis revealed evidence of a stronger effect of increased vmPFC→amygdala inhibition on symptom improvement in individuals who received ketamine (Ep=-0.047, Pp=0.98; **Table S4**).

**Figure 3.**
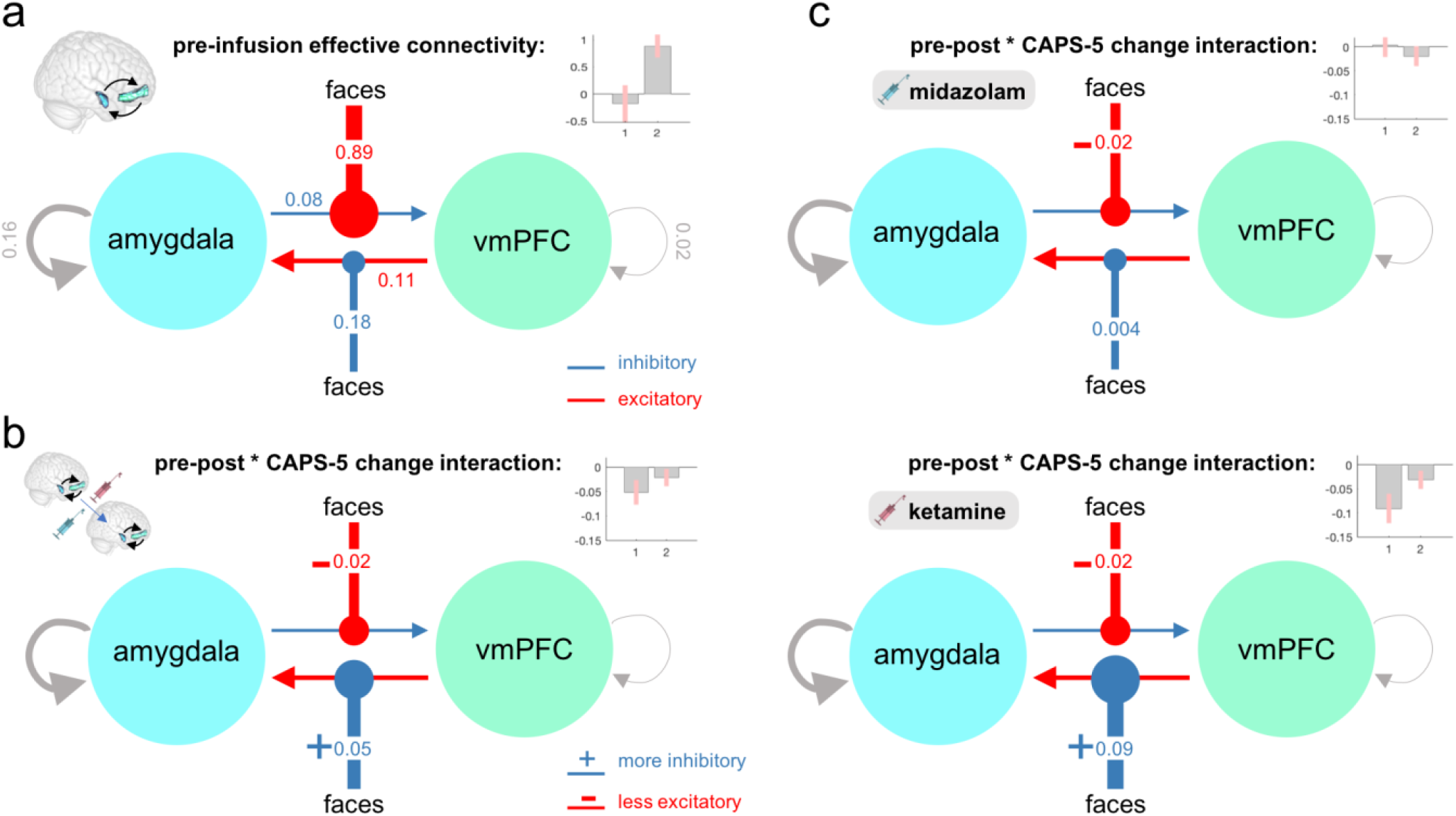
Dynamic causal modelling of task-modulated effective connectivity during the emotional face-processing task. **a** Parametric Empirical Bayes (PEB) analysis of pre-infusion scan data for all participants revealed that, at baseline, the onset of emotional faces shifted the amygdala→vmPFC connection towards an excitation – with only weak evidence for a shift towards top-down inhibition of the AMG by the vmPFC. **b** PEB analysis of pre-post changes in connectivity related to improvement in PTSD symptoms revealed that across all participants, improvement in overall PTSD severity (CAPS-5 total score) was associated with both increased vmPFC inhibition of the amygdala, and decreased amygdala excitation of the vmPFC, during emotional face-viewing. **c** When participants were divided by drug condition, the relationship between PTSD symptom improvement and increased top-down inhibition of the amygdala by the vmPFC during emotional stimuli was only evident in participants who received ketamine, a difference confirmed by passing both models to a 3^rd^ level PEB analysis (more negative parameter estimate for the vmPFC→amygdala connection in the ketamine group, posterior probability=0.98). For all panels, pointed arrowheads represent connections between regions, and circular arrowheads represent modulation of those connections by the experimental condition of interest (emotional faces). Values are rates of change constants in Hz. Insets depict PEB posterior parameter estimates for modulation of connectivity by the listed effect of interest: 1, vmPFC→amygdala; 2, amygdala→vmPFC; error bars represent 90% confidence intervals.

### Baseline prediction of PTSD symptom improvement

A second set of analyses examined whether any of the candidate set of baseline measurements were reliably predictive of symptom improvement following treatment. Across all study participants, the strongest predictor of clinical response (greater PTSD symptom improvement) was lower baseline vmPFC-amygdala connectivity during the emotional face-viewing task (β=-6.03; **Figure S8**). Moderate-to-large effects were also observed for lower baseline rACC BOLD during both emotional face-viewing and emotional conflict regulation tasks (βs=-4.83, -1.30; **Figure 4b**), and in individuals with more distinct representation of fearful vs neutral faces across rACC voxels (β=1.35; **Figure 4c**). Small predictive effects were retained for clinical variables related to baseline clinical severity (pre-infusion CAPS-5 total score, and concurrent use of psychotropic medication or cannabis; βs=0.15, 0.62, 0.56), which likely reflect a regression to the mean effect (tendency for greater improvement at follow-up in individuals with more extreme values at baseline, [60,61]; there was no evidence that baseline vmPFC-amygdala face-related connectivity was related to baseline PTSD severity, *r*=0.04). Other small effects retained in the minimum MSE model are not discussed further here, given the likelihood that these may reflect over-fitting (whole model *r*^2^=0.857).

**Figure 4.**
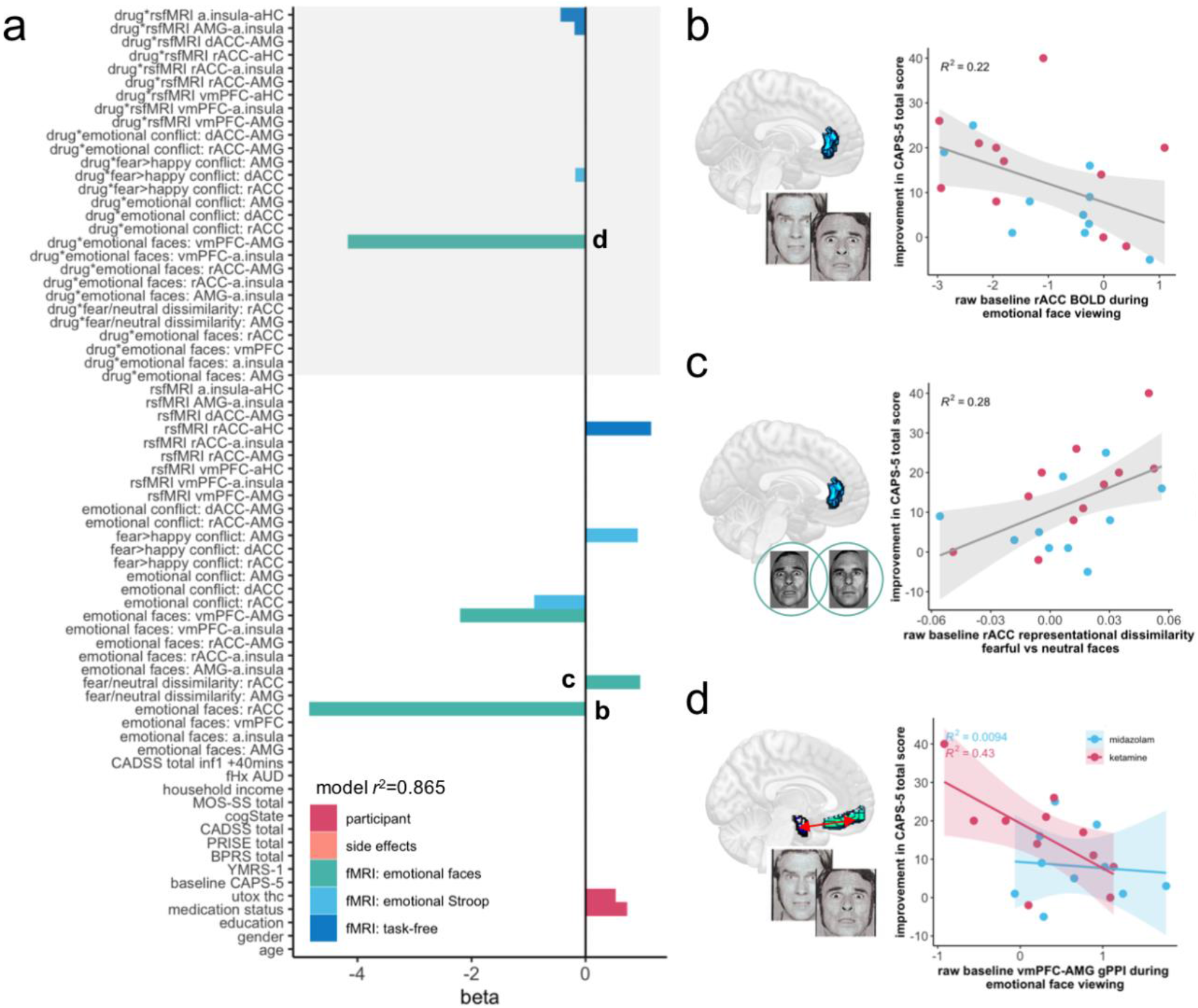
Baseline predictors of PTSD symptom change. **a** Standardized regression coefficient (beta) values for the elastic net model with minimum predictive error for change in PTSD symptoms over the course of treatment in left-out subjects. Imaging measures represent estimates extracted from the baseline (pre-infusion) session. Grey shading highlights interaction terms between imaging measures and received drug identity (ketamine vs midazolam). fHX AUD, family history of alcohol use disorder in first degree relatives; MOS-SS, Medical Outcomes Study Social Support Survey total score; cogState, composite score for executive function derived from the cogState neurocognitive test battery. Lower rACC BOLD during emotional face-viewing (**b**) and increased baseline representational distance between fearful and neutral faces in the rACC (**c**) were retained in the minimum error model as baseline predictors of PTSD symptom improvement across all study participants. **d** Decreased baseline connectivity between the vmPFC and amygdala (AMG) during emotional face-viewing was predictive of symptom improvement across all participants, with evidence of a stronger effect in individuals who went on to receive ketamine (interaction with drug retained in the model with minimum predictive error). Regional BOLD signal values are in arbitrary units. Representational dissimilarity was evaluated using cross-validated linear discriminant contrast estimates, equivalent to the Mahalanobis distance between patterns of response evoked by each kind of stimulus. For generalized psychophysiological interaction (gPPI) functional connectivity estimates, values represent Fisher-transformed correlation coefficients. Bivariate plots represent ‘raw’ imaging measure values (i.e., calculated prior to regression against average within-scan framewise displacement).

When interactions between drug and imaging measures were entered into the model, the interaction between drug and face-related vmPFC-amygdala connectivity was retained (**Figure 4a**), providing evidence of a stronger relationship between baseline connectivity and PTSD symptom improvement in individuals who went on to receive ketamine (β=-4.15, whole model *r*^2^=0.865; **Figure 4d**). When change in MADRS score was included in the model, the interaction between drug and face-related vmPFC-amygdala connectivity was retained (β=-5.21, whole model *r*^2^=0.805; **Figure S9**), indicating some specificity in predicting improvements in cardinal PTSD symptoms, in particular in individuals who went on to receive ketamine.

Exploratory follow-up analysis of baseline effective connectivity data revealed that, across all participants, greater improvement over the course of treatment was associated with lower baseline inhibition of the amygdala by the vmPFC (Ep=0.05, Pp=1.00), and greater baseline excitement of the vmPFC by the amygdala, during viewing of emotional face stimuli (Ep=0.02, Pp=0.98; **Table S5**). Examining PEB results for each drug group separately revealed that the amygdala→vmPFC effect was present in both groups (Ep=0.046, 0.0229, Pp=1.00, 0.99) – but that the relationship between ineffective baseline vmPFC inhibition of the amygdala and treatment response was only evident in individuals who went on to receive ketamine (Ep=0.011, 0.091, Pp=0.71, 0.99; **Table S6**). Formal comparison of drug conditions by passing to a 3^rd^ level PEB analysis revealed evidence of a stronger effect of baseline vmPFC→amygdala connectivity in predicting response in the ketamine group (Ep=0.040, Pp=0.95; **Table S4**). Specifically, participants who responded better to ketamine tended to lack inhibitory modulation of the amygdala by the vmPFC in response to emotional stimuli at baseline (or show abnormal excitatory modulation of this connection; see Discussion); conversely, those who were already able to harness vmPFC inhibition of the amygdala during emotional face-viewing showed less improvement following treatment (**Figure S10**).

## Discussion

Here, we provide preliminary evidence of changes in functional imaging measures of brain activity associated with improvement in overall PTSD symptom severity in individuals undergoing treatment with repeated-dose intravenous ketamine or midazolam. In a sample of individuals with severe, chronic PTSD, the most reliably identified predictor of symptom improvement across models was increased functional connectivity between the amygdala and vmPFC during viewing of emotional face stimuli. Increased emotion-related vmPFC-amygdala coherence was the strongest correlate of symptom change across all subjects – with evidence of a stronger effect in individuals who received ketamine, and some specificity to reduction in cardinal PTSD symptoms (i.e., over and above concomitant decreases in depressive symptoms; **Figure S3, Figure 2, Figure S4**).

This finding is consistent with a body of evidence from previous imaging studies of PTSD and other trauma-exposed groups: specifically, that individuals with PTSD may exhibit hypoactive prefrontal and hyperactive amygdala responses to trauma cues and trauma-unrelated emotional stimuli [13–17]; that differences in prefrontal-amygdala function are implicated in and predictive of resilient outcomes following trauma exposure [62–64]; and that increases in vmPFC-amygdala resting functional connectivity are observed during trials of the current gold standard treatment for PTSD, prolonged exposure therapy [59,65]. Exploratory follow-up analysis via dynamic causal modelling indicated that whilst responders in both drug conditions showed decreased excitatory influence from the amygdala to the vmPFC in response to emotional faces (perhaps representing a common anxiolytic effect of treatment), greater top-down vmPFC inhibition of the amygdala during emotional face viewing was only related to PTSD symptom reduction under ketamine (**Figure 3**). Intriguingly, this pattern of fronto-limbic connectivity change is similar that observed following 10-day administration of the selective serotonin re-uptake inhibitor (S)-citalopram in healthy volunteers [53]. As emotional faces (particularly fearful, angry, and surprised expressions) can be considered social signals of danger – especially in individuals traumatized by interpersonal violence [66] – this could be interpreted as representing decreased threat-responsivity in midazolam and ketamine treatment responders – via both common and drug-specific pathways.

Further, although we present no data here that directly speak to effects of drug treatment on learning, there is a striking overlap between the brain circuitry associated here with response to ketamine, and neural mechanisms identified as central to successful extinction learning in preclinical models and human studies [67–72]. Although the mechanism of action of ketamine in humans is incompletely understood, it has been proposed that it may help remediate mood and stress-related symptoms by opening a ‘window of plasticity’ that promotes un- or re-learning of maladaptive associations that contribute to these symptoms, via increasing neurogenesis in brain regions negatively impacted by chronic stress – in particular in the medial prefrontal cortex [3,4,8,5–7]. Consistent with this hypothesis, preclinical evidence suggests that, under some conditions, administration of ketamine improves fear extinction learning in rodents [73–75]. It is therefore possible that extinction of maladaptive fear responses related to trauma memories may contribute to symptom improvement in individuals who responded to ketamine [7,9]. In trial participants’ own words when discussing their trauma, “I made peace, I could go past it, I could, can let it go”; “Before, talking about it used to make me feel a terrible feeling…[but now] I have to dig out the memory as if from an attic” [2]. This speculative interpretation is consistent with our exploratory finding that changes in vmPFC-amygdala connectivity were most strongly related to improvements in prototypical PTSD symptoms (re-experiencing, anxious arousal, and avoidance), and less strongly related to changes in low mood/motivational symptoms that are also part of PTSD (**Figure S5**). However, the proposal that facilitated extinction learning may be a mechanism underlying response to ketamine for PTSD needs to be explicitly tested in future work.

Finally, the baseline prediction analysis indicated that, across all participants, the strongest predictor of PTSD symptom improvement over the course of treatment was lower baseline vmPFC-amygdala connectivity during emotional face-viewing (**Figure S8**). Lower pre-infusion vmPFC-amygdala coherence in response to faces was particularly associated with response to ketamine, and to improvement in cardinal PTSD symptoms in participants who went on to receive ketamine (**Figure 4, Figure S9**). Exploratory follow-up effective connectivity analysis suggested that greater overall PTSD symptom reduction following ketamine was associated with lower baseline vmPFC inhibition of amygdala during viewing of emotional stimuli (**Figure S10**). Of note, hypofrontal regulation of the amygdala in response to emotional face stimuli has previously been observed via DCM analysis of data from the same task in anxiety disorder samples [55,56], and under task-free conditions in individuals with non-dissociative PTSD [76,77]. Greater response to either treatment was also predicted by lower baseline rACC activity during both emotional face-viewing and emotional conflict regulation tasks (which has previously been interpreted as reflecting poorer regulation of emotional responses in PTSD samples, [30]), and more disparate representation of fearful *vs* neutral faces in the rACC. The latter finding may represent hyper-sensitivity to social signals of threat [78] and/or a signature related to long-term fear learning – as greater representational differentiation between fear and non-fear-associated stimuli in a network of brain regions including the ACC has previously been demonstrated to be predictive of future fear memory retention [79]. Together, these measures may be reflective of a type of PTSD that is more likely to respond to this form of treatment: i.e., one that is primarily related to more abnormal function in brain circuitry subserving extinction learning and threat responsivity – although this hypothesis needs to be confirmed by further investigation.

Strengths of the work presented here include highly novel data (the first presentation to our knowledge of changes in brain function during ketamine treatment for PTSD), collected during a rigorously controlled randomized clinical trial, with detailed clinical and drug side-effects measures available for all participants. We attempted to leverage previous knowledge gained from imaging studies of PTSD by focusing our analysis on an *a priori* target circuit of relevant regions and connections, as specified in a pre-registered analysis plan. Further, the directionality of effects identified here converges with those identified during previous PTSD treatment studies [59,65].

The major limitation of this work stems from this being a small pilot study, as ketamine is still in development as a therapy for PTSD. A small sample size means that our analyses are likely to be underpowered for small-to-moderate effect sizes. Although we should be adequately powered at *N*=21 to detect moderate-to-large effects (*r* of 0.55 and above with 80% power, see Supplementary Material: bivariate correlation estimate between PTSD symptom improvement and change in vmPFC-amygdala connectivity, *r*=0.54; for vmPFC-amygdala baseline connectivity, *r*=0.51), it is important to note that large effect sizes in small samples may reflect inflated estimates of brain-behaviour associations [80,81]. We urge particular caution when interpreting results from emotion regulation (face Stroop) task, given almost 20% imputed data for this measure. Further, although we attempted to guard against over-fitting by selecting regression penalization parameters via cross-validation, this is not a panacea – particularly at smaller sample sizes, where the error in estimating predictive accuracy in left-out subjects may be relatively large [82]. Recent work has also highlighted that mean univariate BOLD measures derived from emotional processing tasks (including the emotional face-viewing task employed here) may exhibit poor within-subject reliability, particularly for subcortical regions [83–85]. Whilst there is some evidence that reliability may be greater for both PPI functional connectivity estimates [86] and multivariate similarity measures [87,88], measurement reliability fundamentally limits the ability to detect true between-subjects effects in our data. Overall, we believe that our findings should be considered as preliminary, and – rather than a representing a definitive account of brain changes accompanying or predictive of symptom improvement following ketamine for PTSD – as a springboard for hypothesis-testing in future independent datasets.

In summary, improvement in PTSD symptom severity over the course of treatment was related to connectivity changes between regions previously identified as showing abnormal activity in PTSD (specifically, the vmPFC, amygdala, d/rACC, and anterior insula). These changes were primarily observed during processing of socio-emotional stimuli (ambiguous/neutral and negatively valenced emotional faces), which can be considered as threat-related signals in individuals traumatized by interpersonal violence. We provide preliminary evidence for a drug-specific mechanism underlying vmPFC-amygdala connectivity changes following treatment: i.e., that improvement under both midazolam and ketamine was related to decreased amygdala→vmPFC excitation during processing of socio-emotional stimuli, but that only improvement under ketamine was associated with increased top-down inhibition of the amygdala by the vmPFC under these conditions. We provisionally propose that down-regulation of threat responses and/or enhancement of fear extinction learning might contribute to decreases in the severity of prototypical PTSD symptoms following ketamine – effects that may be supported by increased neurogenesis in the medial prefrontal cortex. This may help explain the greater rate of response, and longer time to relapse, in individuals who received ketamine compared to the anxiolytic midazolam [2]. Further, this speculative interpretation is in line with increasing evidence that successful treatments for mood and stress-related disorders are both pharmacological and psychological in nature: i.e., that effective drug treatments appear to ‘open a window’ for re-learning of ingrained cognitive biases or maladaptive fear memories that contribute to symptom maintenance [89–91,9]. Future work could explore the potential of combining ketamine with adjunct treatments that directly target extinction learning (e.g., psychological therapies with an element of imagined re-exposure), in order to test if this might improve response rates or result in longer maintenance of treatment response for this chronic, debilitating disorder [8,18].

## Data Availability

Data associated with this manuscript is not currently publicly available, as permission for public sharing was not sought at the time of participant consent.

https://osf.io/8bewv/

## Funding and Disclosures

This work was supported by an Independent Investigator Award from the Brain and Behavior Research Foundation to AF (grant number 23144), Technology Development Awards from Mount Sinai Innovation Partners and the Mount Sinai i3 Accelerator, and a donation from Mr. Gerald Greenwald and Mrs. Glenda Greenwald. Additional funding was provided by the Ehrenkranz Laboratory for Human Resilience, a component of the Depression and Anxiety Center for Discovery and Treatment at Icahn School of Medicine at Mount Sinai.

Drs. Feder and Charney are named co-inventors on a patent application in the US, and several issued patents outside the US, filed by the Icahn School of Medicine at Mount Sinai (ISMMS) related to the use of ketamine for the treatment of PTSD. This intellectual property has not been licensed. Dr. Jha has received contract research grants from Acadia Pharmaceuticals and Janssen Research & Development, educational grant to serve as Section Editor of the Psychiatry & Behavioral Health Learning Network, consultant fees from Eleusis Therapeutics US, Inc, and honoraria for CME presentations from North American Center for Continuing Medical Education and Global Medical Education. Dr. K. Collins is a paid independent rater for Medavante-Prophase. In the past 5 years, Dr. Murrough has provided consultation services and/or served on advisory boards for Allergan, Boehreinger Ingelheim, Clexio Biosciences, Fortress Biotech, FSV7, Global Medical Education (GME), Novartis, Otsuka, Sage Therapeutics, and Engrail Therapeutics. Dr. Murrough is named on a patent pending for neuropeptide Y as a treatment for mood and anxiety disorders and on a patent pending for the use of KCNQ channel openers to treat depression and related conditions. The ISMMS (employer of Dr. Murrough) is named on a patent and has entered into a licensing agreement and will receive payments related to the use of ketamine or esketamine for the treatment of depression. The ISMMS Medicine is also named on a patent related to the use of ketamine for the treatment of PTSD. Dr. Murrough is not named on these patents and will not receive any payments. Dr. Charney is named as co-inventor on patents filed by the ISMMS relating to the treatment for treatment-resistant depression, suicidal ideation and other disorders. ISMMS has entered into a licensing agreement with Janssen Pharmaceuticals, Inc. and it has and will receive payments from Janssen under the license agreement related to these patents for the treatment of treatment-resistant depression and suicidal ideation. Consistent with the ISMMS Faculty Handbook (the medical school policy), Dr. Charney is entitled to a portion of the payments received by the ISMMS. Since SPRAVATO has received regulatory approval for treatment-resistant depression, ISMMS and thus, through the ISMMS, Dr. Charney, will be entitled to additional payments, beyond those already received, under the license agreement. Dr. Charney is a named co-inventor on several patents filed by ISMMS for a cognitive training intervention to treat depression and related psychiatric disorders. The ISMMS has entered into a licensing agreement with Click Therapeutics, Inc. and has and will receive payments related to the use of this cognitive training intervention for the treatment of psychiatric disorders. In accordance with the ISMMS Faculty Handbook, Dr. Charney has received a portion of these payments and is entitled to a portion of any additional payments that the medical school might receive from this license with Click Therapeutics. Dr. Charney is a named co-inventor on a patent application filed by the ISMMS for the use of intranasally administered Neuropeptide Y (NPY) for the treatment of mood and anxiety disorders. This intellectual property has not been licensed. The other authors declare no conflicts of interest.

## Author contributions

Contribution to the conception or design of the work: MKJ, LMS, DSC, JWM, AF. Contribution to the acquisition, analysis, or interpretation of data for the work: AN, SBR, ABC, SC, SRH, MK, MC, KAC, AMG, JB, JMW, AF. Drafting the work or revising it critically for important intellectual content: AN, SC, MKJ, LMS, JMW, AF. Agreement to be accountable for all aspects of the work in ensuring that questions related to the accuracy or integrity of any part of the work are appropriately investigated and resolved: AN, AF. Final approval of the version to be published: All authors.

## Acknowledgments

This work was supported in part through the computational resources and staff expertise provided by Scientific Computing at the Icahn School of Medicine at Mount Sinai.

## Notes

### Clinical Trial

NCT02397889

### Clinical Protocols

https://osf.io/8bewv/

### Author Declarations

The study received ethical approval from the Institutional Review Board at the Icahn School of Medicine at Mount Sinai.

