## Supplementary Material for "Neuroimaging correlates and predictors of response to repeated-dose intravenous ketamine in PTSD: preliminary evidence"

### Supplementary Methods

#### Participants

Full inclusion and exclusion criteria for the randomized controlled trial imaging data was collected during are available at https://clinicaltrials.gov/ct2/show/NCT02397889. Major inclusion criteria for the trial were being aged 18-70, meeting full DSM-5 criteria for PTSD as determined by the Structured Clinical Interview for DSM-5 (SCID-5; [1]), past-month CAPS-5 total score at screening ≥ 30 (several points into the moderate severity range), and ability to give informed consent. Exclusion criteria included serious unstable medical illnesses, history of psychosis, substance use disorder within the last 3 months, or treatment with a long-acting benzodiazepine or opioid drug within the two weeks prior to randomization. Exclusion criteria for the MRI sessions were claustrophobia, any trauma or surgery which may have left magnetic material in the body, presence of magnetic implants or pacemakers, and inability to lie still for one hour or more without discomfort.

#### Medication status and urine toxicology

Participants for the wider clinical trial were permitted to be on stable doses (≥3 months) of certain psychotropic medications (e.g., antidepressants, see above). Individuals who reported current use of marijuana or cannabis derivatives were also not excluded (absent meeting criteria for current substance use disorder), due to high prevalence of self-medication in people with PTSD. Urine toxicology testing was performed prior to both imaging sessions. Per the analysis plan, participants positive for tetrahydrocannabinol (THC) were not excluded from the imaging analysis; however, current use of a psychotropic medication and/or positive test for THC on either scan session were recorded and entered into the analysis.

#### Drug side-effects measures

Several clinician- and patient-rated side-effects measures were included in the analysis. Specifically, these were the Clinician-Administered Dissociative States Scale (CADSS), a measure of dissociative drug effects [2]; four items from the Brief Psychiatric Rating Scale (BPRS) probing psychotomimetic symptoms (17); and a single item from the Young Mania Rating Scale (YMRS), indexing elevated mood [4]. These three measures have previously been shown to be sensitive to the psychoactive effects of ketamine, and to differentiate between intravenous ketamine and midazolam – with peak differential responses at 40 minutes post-infusion [5,6]. For each measure, average drug-related effects across infusions were calculated as mean scores at 40 minutes post-infusion minus mean scores pre-infusion. Data from the Patient Rated Inventory of Side-Effects (PRISE; 19) was also included in the analysis, as this measure is sensitive to effects of both ketamine and midazolam (e.g., nausea, fatigue) [5,6]. For PRISE data, scores were summed across all somatic domains, and mean drug-related effects were calculated as average scores at the end of each infusion session, minus baseline.

#### Other baseline demographic and clinical measures

Available demographic information included age, self-reported gender identity, self-reported race and ethnicity, self-reported education level, and self-reported income (see [8] for full details). Low-frequency gender identities were collapsed into two categories (male, and female/other or non-binary), in order to preserve participant privacy and enable estimable categorical effects.

Total scores of the Medical Outcomes Study Social Support Survey (MOS-SS), a self-report measure which assesses perceived levels of social support [9], was included in the baseline prediction analysis, as social support has previously been shown to play an important role in resilience to psychopathology following trauma [10]. Participants also completed a computerized battery of executive function and working memory tests (CogState), that has been shown to be sensitive to cognitive impairment [11]. Performance across tasks on a primary index measure was z scored across participants, then averaged to yield a composite score for cognitive function, as per the CogState analysis guide. Composite cognitive function scores were included in the baseline prediction model, given evidence that neurocognitive function may be associated with PTSD symptom severity in traumatized samples [12,13]. We also recorded self-reported family history of alcohol use disorder in first degree relatives, as this has previously been linked to response to single-dose intravenous ketamine in depression [14]. CADSS score changes during the first infusion were included in the prediction analysis as pseudo-baseline measure, on the basis of evidence that initial dissociative effects may be associated with rapid-onset antidepressant effects of ketamine [15].

#### Neuroimaging data

##### Preprocessing

Image preprocessing was performed using fMRIPprep 1.1.4 [16,17], which is based on Nipype 1.1.1 [18,19].

**Anatomical data preprocessing.** A total of two T1-weighted (T1w) images were available in the input dataset (across sessions). Both images were corrected for intensity non-uniformity (INU) using N4BiasFieldCorrection [20], ANTs 2.2.0. A T1w-reference map was computed after registration of the 2 T1w images (after INU-correction) using mri_robust_template (FreeSurfer 6.0.1) [21]. The T1w-reference was then skull-stripped using antsBrainExtraction.sh (ANTs 2.2.0), using OASIS as the target template. Spatial normalization to the ICBM 152 Nonlinear Asymmetrical template version 2009c [22] was performed through nonlinear registration with antsRegistration (ANTs 2.2.0) [23], using brain-extracted versions of both T1w volume and template. Brain tissue segmentation of cerebrospinal fluid (CSF), white matter (WM) and grey matter (GM) was performed on the brain-extracted T1w using fast (FSL 5.0.9) [24].

**Functional data preprocessing.** For each of the six BOLD runs available per subject (three tasks across two sessions), the following preprocessing was performed. First, a reference volume was generated using a custom methodology of fMRIPrep. Head-motion parameters with respect to the BOLD reference (transformation matrices, and six corresponding rotation and translation parameters) were estimated before any spatiotemporal filtering using mcflirt (FSL 5.0.9) [25]. BOLD runs were then slice-time corrected using 3dTshift from AFNI. The BOLD slice-time corrected time-series were resampled onto their original, native space by applying a single, composite transform to correct for head-motion and susceptibility distortions. These resampled BOLD time-series will be referred to as preprocessed BOLD. The BOLD reference was then co-registered to the T1w reference using flirt (FSL 5.0.9) [26] with a boundary-based registration cost function [27]. Co-registration was configured with nine degrees of freedom to account for distortions remaining in the BOLD reference.

Automatic removal of motion artifacts using independent components analysis (ICA-AROMA) [28] was performed on the preprocessed BOLD time-series after a spatial smoothing with an isotropic, Gaussian kernel of 6mm FWHM (full-width half-maximum). Corresponding "non-aggressively" denoised runs were produced after such smoothing. The BOLD time-series were resampled to MNI152NLin2009cAsym standard space, generating a preprocessed BOLD run in MNI152NLin2009cAsym space. Several confounding time-series were calculated based on the preprocessed BOLD: framewise displacement (FD), DVARS and three region-wise global signals. FD and DVARS are calculated for each functional run, both using their implementations in Nipype (following the definitions by Power et al, [29]). The three global signals were extracted within the CSF, the WM, and the whole-brain masks. Additionally, a set of physiological regressors were extracted to allow for component-based noise correction (CompCor, [30]). Principal components were estimated after high-pass filtering the preprocessed BOLD time-series (using a discrete cosine filter with 128s cut-off) for the two CompCor variants: temporal (tCompCor) and anatomical (aCompCor). For aCompCor, six components were calculated within the intersection of the aforementioned mask and the union of CSF and WM masks calculated in T1w space, after their projection to the native space of each functional run (using the inverse BOLD-to-T1w transformation).

All resamplings were performed with a single interpolation step by composing all the pertinent transformations (i.e. head-motion transform matrices, susceptibility distortion correction when available, and co-registrations to anatomical and template spaces). Gridded (volumetric) resamplings were performed using antsApplyTransforms (ANTs), configured with Lanczos interpolation to minimize the smoothing effects of other kernels [31]. Non-gridded (surface) resamplings were performed using mri_vol2surf (FreeSurfer).

**First level analysis models**

Full specification of first-level models is available at the OSF project page (https://osf.io/8bewv/). In order to account for potentially incomplete ICA-AROMA removal of motion-related effects [32,33], and in line with the original approach of Pruim et al. [28,34], several confound regressors generated by fMRIprep were also included in all first-level models: specifically, mean time series extracted from CSF and WM masks, six components estimating spatially-coherent noise in these tissues (aCompCor) [30,35], and additional outlier regressors for any volume with framewise displacement (how much the head changed position from one frame to the next)>0.5mm or DVARS (how much image intensity changed from one frame to another)>0.5% [36]. (Although these regressors are calculated by fmriprep prior to non-aggressive ICA-AROMA denoising, simulation evidence suggests this additional denoising step is equivalent or even slightly beneficial compared to inclusion quantities calculated following ICA-AROMA; see https://github.com/nipreps/fmriprep-notebooks).

**Emotional face-processing task.** Two-hundred functional volumes were collected, including 5 dummy volumes at the start of the task that were discarded in order to allow for T1 equilibrium effects. Onsets of shapes and emotional faces were modelled as boxcar functions for the duration of stimulus presentation (4000ms). Separate regressors were included for the onset of each of the 5 types of emotional face. A further regressor was included for trials on which an error was made (incorrect matching shape or face stimulus selected), in order to account for potential attentional lapses. The main contrast of interest submitted to further analysis for mean univariate BOLD and functional connectivity analysis was all faces > shapes trials.

**Emotional conflict regulation (face Stroop) task**. Two-hundred and eighteen functional volumes were collected, including 5 dummy volumes. The first level model included separate onsets for congruent, incongruent, and baseline trials, divided by target face emotion (fearful or happy). These 6 regressors were modelled as boxcar functions for the duration of stimulus presentation (2000ms). A further regressor was included for trials on which an error was made (incorrect facial emotion selected). The main contrast of interest submitted to further analysis was incongruent > congruent trials. A subset of measures also examined the fear incongruent > happy incongruent contrast (incongruent trials where the target was an afraid face *vs* incongruent trials where the target was a happy face), in order to identify neural processing during emotional conflict regulation specific to negative emotions or social signals of threat.

**Task-free (resting state) scan**. 300 functional volumes were collected during the 12-min task-free scan. Regressors of no interest were the same as for the task-based scans (see above). Additional band-pass filtering was not carried out on the basis that ICA-AROMA already performs a form of flexible high-pass filtering [28], which may preserve more signal than standard band-pass filtering [34], and informed by benchmarking evidence that suggests that ICA-AROMA plus additional nuisance regression based on expansions of CSF and WM time-series performs well on task-free data [37].

**Derivation of imaging measures**

**Mask definition for functionally-defined regions of interest (ROIs)**. Masks for ROIs that are not well-defined anatomically (vmPFC, rACC, dACC, aHC) were generated from meta-analytic consensus maps using Neurosynth [38]. Specifically, association maps (maps depicting whether activation in a given voxel occurred more consistently for studies in the literature that mention a particular term, compared to studies that don't, thresholded using a false discovery rate of 0.01) were generated for the search terms 'rostral anterior cingulate', 'dorsal anterior cingulate', 'ventromedial prefrontal cortex', and 'anterior hippocampus'. These statistical images were then further thresholded ($z>1$), masked by relevant wider anatomical region (ACC, mPFC, and HC, respectively), and binarized using the SPM12 Imcalc utility. rACC/dACC masks were further constrained to be mutually exclusive by extracting one from each other, in order to remove any overlapping voxels. Of note, the rACC ROI used here encompasses the pregenual region usually implicated in studies of

PTSD, as opposed to the subgenual anterior region (BA25 and PL32) often implicated in studies of depression. Mask images are available at the OSF project page (https://osf.io/8bewv/).

**Mean (univariate) signal extraction**. Mean BOLD signal across ROI masks were extracted from the contrast of interest for each task using MarsBaR, version 0.44 [39].

**Functional connectivity analysis**. Task-specific ROI-ROI covariances were modelled using generalized psychophysiological interaction (gPPI) analysis, as implemented in the CONN toolbox, version 18.b [40]. Denoising was via the approach described previously (i.e., non-aggressive ICA-AROMA within fMRIprep, followed by first-level analysis models in SPM12 that included mean time series in WM and CSF masks, plus six aCompCor components describing spatially-coherent noise in these tissues, and outlier regressors for volumes with FD>0.5mm or DVARS>0.5%). Average time-series were then extracted for each ROI, using subject-specific GM masks. PPI analysis evaluates how well the *interaction* or product between a task-condition (psychological) vector and BOLD (physiological) time-series in a seed ROI explains the BOLD time-series in a target ROI (i.e., identifies relative bivariate correlation or connectivity changes associated with onset of a specific experimental condition). In gPPI, the interaction factors from all conditions of interest are considered simultaneously during estimation, in order to better account for between-condition overlap [41]. Here, ROI-ROI gPPI functional connectivities for each task-based scan were estimated via bivariate regression of target against seed*task interaction time-series, with HRF-convolved weighting of the interaction terms. For the task-free scan, this analysis was equivalent to a weighted GLM examining a single regressor encompassing the entire scan duration.

**Multivariate representational similarity analysis**. In order to assess multivariate representation of emotional faces, multivariate similarity metrics were extracted for the face-processing task using the RSA toolbox [42]. Time-series data were extracted from first level model for each region of interest, multivariately noise normalized, and separated into two partitions (alternating trials). The cross-validated linear discriminant contrast (Mahalanobis distance) between multivoxel representations of different emotional face stimuli, averaged across all presentations, was then calculated. This (dis)similarity metric was chosen as it is unbiased by noise (has a meaningful zero point), and is more reliable than estimates of similarity based on correlation coefficients [43].

**Statistical analysis**

**Sample size.** As intravenous ketamine is still under development as a therapy for PTSD, this is a small, pilot study. A small sample size means that our analyses are likely underpowered for small-to-moderate size effects, although power should be adequate to detect moderate-to-large (potentially most clinically relevant) effects (for a simple bivariate relationship at $N$=21, we should be able to detect an $r$ of 0.55 or above for alpha=0.05, two-tailed, and 80% power).

**Missing data.** As noted at time of pre-registration, one participant who completed the baseline scan and all clinical trial procedures declined to complete a post-infusion scan, and face Stroop task data were not collected for $N$=3 participants. In addition, one participant declined to provide information about household income, and one participant did not complete the baseline cognitive test battery. Per our analysis plan, missing data was dealt with via multiple imputation, as implemented in the R package mice [44]. This method replaces missing values with plausible simulated data which leave the mean and variance of the distribution for each variable unchanged.

**Deviations from pre-registered analysis.** Diffusion-weighted images were also collected, but unfortunately we were not able to analyse these as planned (i.e., via anatomically-constrained tractography between our target ROIs; [45]) due to incompletely collected distortion-correction (field-map) images. These data are therefore not further discussed here. It was further decided to include mean dissociative symptoms during infusion sessions in all analysis models (as well as manic and psychotomimetic side-effects), in order to account for potential functional unblinding of participants in the ketamine group.

**Exploratory follow-up analyses**

**Effective connectivity estimation via dynamic causal modelling.** In order to gain insight into the directionality of the functional connectivity effects identified in the main analysis (which simply evaluates symmetrical covariances between pairs of ROIs), follow-up effective connectivity analysis of task fMRI data was carried out via Dynamic Causal Modelling (DCM) [46,47]. Briefly, DCM involves specifying a biologically-informed ('forward') model of how activity in one brain region affects activity in another, and how this might be reflected in measured BOLD output. For a given model, the probability of the observed BOLD signal data can then be estimated via model inversion.

For each subject and session, time-series were extracted from voxels within a 6mm sphere around the subject-specific peak for the contrast of interest, within each ROI mask. In order to reduce noise, time-series were first adjusted for nuisance effects (those not covered by a general 'all effects of interest' F contrast that included onset of all trial types), and only voxels within each sphere which met a liberal threshold of $p<0.05$ for the contrast of interest were extracted (this threshold is not used to determine statistical significance of any results, but rather represents a standard pre-processing step to focus the analysis on voxels most related to the task of interest for each participant; [47]). Time-series were then summarized using the first eigenvariate across voxels, and summarized time-series used to construct Dynamic Causal Models (DCMs) in SPM12 [48]. Based on previous DCM analyses of the emotional face-processing task ([49–52]), trial onset (all conditions) was allowed to drive input to each ROI, and onset of the condition of interest (e.g., emotional faces) was allowed to modulate connections between ROIs. Individual DCMs were estimated via Variational Laplace inversion using default shrinkage priors. Estimated DCMs were submitted to further analysis via hierarchical Parametric Empirical Bayes (PEB) [53,54], in order to characterize intersubject variability in task-modulated connectivity: i.e., identify changes in connectivity that scaled with PTSD symptom improvement (see [55]).

Pre-infusion imaging sessions DCMs for all subjects were first submitted to a 2$^{nd}$ level PEB model, in order to identify mean baseline task-related modulation of effective connectivity between ROIs. In a second analysis, changes in pre *vs* post task-related modulation of connectivity associated with improvements in PTSD symptom severity were investigated by inspecting the results of a 2$^{nd}$ level PEB model which included a regressor representing the interaction of change in CAPS-5 total scores and time point, controlling for main effects of time (session) and CAPS-5 score changes (all regressors were mean-centered). The existence of an interaction between these changes and the factor of drug was investigated by constructing 2nd level PEB models as described above, separately for individuals who received midazolam and ketamine, and then passing these to a 3rd level PEB model to test for differences between groups (i.e., the presence of pre-post*CAPS-5 change*drug interactions). Finally, in order to investigate associations between baseline effective connectivity estimates and response to treatment, pre-infusion DCM models were analyzed via a 2nd level PEB with a mean-centered covariate of CAPS-5 total score improvement (both over all subjects, and separately via drug group). Differences in baseline response prediction according to received drug identity were probed by passing these results to a 3rd level PEB, as for the pre-post change analysis. Reported effective connectivity parameters are rates of change in Hz (i.e., for each connection A→B, the rate of change in region B effected by region A; [47]).

#### Supplementary Results

**Change in symptoms following drug treatment in the imaging sample.** Although examining differences in efficacy between repeated-dose ketamine and midazolam for PTSD was not a primary aim of this analysis (please see [56]), repeated-measures ANOVA was used to analyse if improvement in clinical outcome measures examined here (PTSD and depression symptoms) differed between those who received ketamine and midazolam in the imaging sub-sample. Specifically, repeated-measures ANOVA with the within-subjects factors of session (baseline vs outcome visit) and measure (CAPS-5 and MADRS total scores), and the between-subjects factor of drug (ketamine vs midazolam) revealed a significant interaction between factors of session and drug ($F_{1,57}$=6.58, $p$=0.013; **Figure 1b**). Follow-up contrasts revealed that, across measures, symptom scores did not differ at baseline $t_{26}$=0.17, $p$=0.86), but were lower at outcome in individuals who received ketamine compared to midazolam ($t_{26}$=2.2, $p$=0.04). At this sample size, there was no evidence of an interaction between session, drug, and measure (i.e., differential effect of ketamine on PTSD vs depression total symptom scores; $F_{1,57}$<1, $p$>0.5).

**Imaging task behavioural performance across pre and post-infusion scan sessions**. Although differences in behavioural performance (response accuracy and RTs) on the imaging tasks did not form part of our primary study hypothesis, we analyse them here for completeness. For the emotional face-processing task, there was no evidence of an effect of session (pre- *vs* post-infusions) or drug (ketamine *vs* midazolam) on face-matching accuracy or average response time measures (all $p$>0.2, repeated-measures ANOVAs with within-subjects factor of session, and between-subjects factor of drug; **Figure S1a,b**). For the emotional conflict regulation task, there was marginal evidence for an interaction between drug and session on overall response accuracy ($F_{1,47}$=4.28, $p$=0.04; repeated-measures ANOVA with within-subjects factors of task condition [incongruent vs congruent] and session, and between-subjects factor of drug; no evidence of a main effect of task condition on response accuracy, $p$>0.2). This appeared to be driven by a tendency towards lower accuracy on the post-infusion session in participants who had received midazolam (**Figure S1c**). There was no evidence of an effect of session or drug on response times (all $p$>0.1; which again, did not differ by task condition, $p>0.3$; **Figure S1d**). Notably, for both tasks, performance (response accuracy) was close to ceiling level for both pre- and post- infusion imaging sessions.

**Distribution of clinical outcome measures and collinearity of imaging measures.** The distribution of the target variable for the main analysis (pre-post changes in CAPS-5 total scores) did not differ significantly from normal ($p>0.5$, Shapiro-Wilk test). Distributions of other clinical and side-effects variables are visualized as histograms in **Table S1**. Correlations across variables included in the correlation of symptom change and baseline prediction analyses are depicted in **Figure S2** and **Figure S7**, respectively.

| Variable | Values | Histogram |
|---|---|---|
| Change in past-week CAPS-5 total score (baseline *vs* outcome visits) | mean (SD) : 12.2 (11.2)<br>min < med < max: -5 < 11 < 40<br>IQR (CV) : 17 (0.9) | 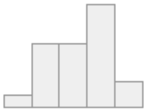 |
| Change in past-week MADRS total score (baseline *vs* outcome visits) | mean (SD) : 10.0 (9)<br>min < med < max: -6 < 8 < 28<br>IQR (CV) : 9 (0.9) | 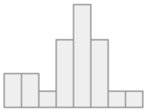 |
| Mean change in CADSS total score across infusion sessions (40 minutes post-infusion *vs* pre-infusion) | mean (SD) : 3.2 (5)<br>min < med < max: -0.3 < 1 < 19.7<br>IQR (CV) : 3 (1.6) | 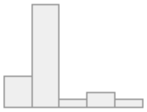 |
| Mean change in BPRS score across infusion sessions (40 minutes post-infusion *vs* pre-infusion) | mean (SD) : 0.2 (0.2)<br>min < med < max: 0 < 0.2 < 1<br>IQR (CV) : 0 (1) | 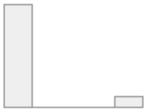 |
| Mean change in YMRS-1 score across infusion sessions (40 minutes post-infusion *vs* pre-infusion) | mean (SD) : 0.1 (0.2)<br>min < med < max: 0 < 0 < 0.8<br>IQR (CV) : 0 (3.3) | 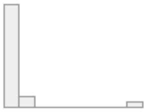 |
| Mean change in PRISE total score across infusion sessions (240 minutes post-infusion *vs* pre-infusion) | mean (SD) : 1.3 (0.7)<br>min < med < max: 0.5 < 1.2 < 3.3<br>IQR (CV) : 0.9 (0.6) | 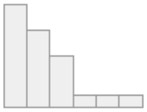 |

**Table S1**. **Summary statistics and distributions of clinical and side-effects measures used in the analysis**. CAPS-5, Clinician-Administered PTSD Scale for DSM-5; MADRS, Montgomery-Åsberg Depression Rating Scale; CADSS, Clinician-Administered Dissociative States Scale; BPRS, four items from the Brief Psychiatric Rating Scale probing psychotomimetic symptoms; YMRS, a single item from the Young Mania Rating Scale indexing elevated mood; PRISE, Patient Rated Inventory of Side-Effects (total score calculated by summing across all somatic domains). Table generated using the R package summarytools [57].

| | Ep | 90% CI | Pp |
|---|---|---|---|
| **extrinsic connectivity estimates (mean)** | | | |
| vmPFC to vmPFC (self-inhibition) | 0.02 | -0.05 – 0.09 | 0.66 |
| vmPFC to amygdala | 0.11 | 0.05 – 0.17 | 1.00 |
| amygdala to vmPFC | -0.08 | -0.12 – -0.04 | 1.00 |
| amygdala to amygdala (self-inhibition) | 0.16 | 0.10 – 0.22 | 1.00 |
| **modulation by emotional faces (mean)** | | | |
| vmPFC to amygdala | -0.18 | -0.52 – 0.16 | 0.81 |
| amygdala to vmPFC | 0.89 | 0.67 – 1.10 | 1.00 |

**Table S2. 2$^{nd}$ level PEB analysis of baseline (pre-infusion) emotional face-viewing task data (mean effects across all participants)**. All network connections (extrinsic and task-related modulation of connections) were modelled as random-effects across participants. Positive modulatory effects represent a shift towards greater excitation or less inhibition, and negative effects represent a shift towards lower excitation or greater inhibition, during emotional face-viewing. vmPFC, ventromedial prefrontal cortex; Ep, posterior estimate for (modulation of) effective connectivity; Pp, posterior probability.

| | Ep | 90% CI | Pp |
|---|---|---|---|
| **modulation by emotional faces (pre-post change*CAPS-5 improvement): all participants** | | | |
| vmPFC to amygdala | -0.052 | -0.077 – -0.026 | 1.00 |
| amygdala to vmPFC | -0.022 | -0.039 – -0.004 | 0.98 |
| **modulation by emotional faces (pre-post change*CAPS-5 improvement): midazolam** | | | |
| vmPFC to amygdala | 0.004 | -0.02 – 0.03 | 0.60 |
| amygdala to vmPFC | -0.019 | -0.04 – 0.001 | 0.94 |
| **modulation by emotional faces (pre-post change*CAPS-5 improvement): ketamine** | | | |
| vmPFC to amygdala | -0.091 | -0.12 – -0.06 | 1.00 |
| amygdala to vmPFC | -0.031 | -0.05 – -0.01 | 1.00 |

**Table S3. 2nd level PEB analysis of pre and post-infusion emotional face-viewing task data for all participants (interaction between pre-post changes and improvement in CAPS-5 total score), across all subjects, and separately for individuals received midazolam and ketamine**. Task-related modulation of connections were modelled as random-effects across participants. Positive effects represent greater excitation or less inhibition, and negative effects represent lower excitation or greater inhibition, post vs pre-infusion, in individuals who showed greater improvement in PTSD symptoms over the course of treatment. vmPFC, ventromedial prefrontal cortex; Ep, posterior estimate for modulation of effective connectivity; Pp, posterior probability.

| | Ep | 90% CI | Pp |
|---|---|---|---|
| **modulation by emotional faces (pre-post change*CAPS-5 improvement*drug)** | | | |
| vmPFC to amygdala | -0.047 | -0.085 – -0.001 | 0.98 |
| amygdala to vmPFC | -0.006 | -0.040 – 0.029 | 0.61 |

**Table S4. 3rd level PEB analysis of pre and post-infusion emotional face-viewing task data (interaction between pre-post changes, improvement in CAPS-5 total score, and received drug identity).** Task-related modulation of connections were modelled as random-effects across participants. Negative effects represent lower excitation or greater inhibition, post vs pre-infusion, in individuals who showed greater improvement in PTSD symptoms following ketamine (compared to midazolam). vmPFC, ventromedial prefrontal cortex; Ep, posterior estimate for modulation of effective connectivity; Pp, posterior probability.

| | Ep | 90% CI | Pp |
|---|---|---|---|
| **modulation by emotional faces (relationship to CAPS-5 improvement): all participants** | | | |
| vmPFC to amygdala | 0.050 | 0.024 – 0.076 | 1.00 |
| amygdala to vmPFC | 0.020 | 0.004 – 0.037 | 0.98 |
| **modulation by emotional faces (relationship to CAPS-5 improvement): midazolam** | | | |
| vmPFC to amygdala | 0.011 | -0.02 – 0.04 | 0.71 |
| amygdala to vmPFC | 0.046 | 0.02 – 0.078 | 1.00 |
| **modulation by emotional faces (relationship to CAPS-5 improvement): ketamine** | | | |
| vmPFC to amygdala | 0.091 | 0.060 – 0.12 | 1.00 |
| amygdala to vmPFC | 0.029 | 0.009 – 0.048 | 0.99 |

**Table S5. 2nd level PEB analysis of baseline(pre-infusion) emotional face-viewing task data for all participants, identifying covariates of future changes in CAPS-5 total score (predictors of response), across all subjects, and separately for individuals who went on to receive midazolam and ketamine**. Task-related modulation of connections were modelled as random-effects across participants. Positive effects represent greater excitation or less inhibition, and negative effects represent lower excitation or greater inhibition, pre-infusion, in individuals who showed greater improvement in PTSD symptoms over the course of treatment. vmPFC, ventromedial prefrontal cortex; Ep, posterior estimate for modulation of effective connectivity; Pp, posterior probability.

| | Ep | 90% CI | Pp |
|---|---|---|---|
| **modulation by emotional faces (CAPS-5 improvement*drug)** | | | |
| vmPFC to amygdala | 0.040 | 0.001 – 0.08 | 0.95 |
| amygdala to vmPFC | -0.009 | -0.044 – 0.027 | 0.66 |

**Table S6. 3rd level PEB analysis of baseline (pre-infusion) emotional face-viewing task data (interaction between future improvement in CAPS-5 total score, and received drug identity).** Task-related modulation of connections were modelled as random-effects across participants. Negative effects represent lower excitation or greater inhibition, pre-infusion, in individuals who showed greater improvement in PTSD symptoms following ketamine (compared to midazolam). vmPFC, ventromedial prefrontal cortex; Ep, posterior estimate for modulation of effective connectivity; Pp, posterior probability.

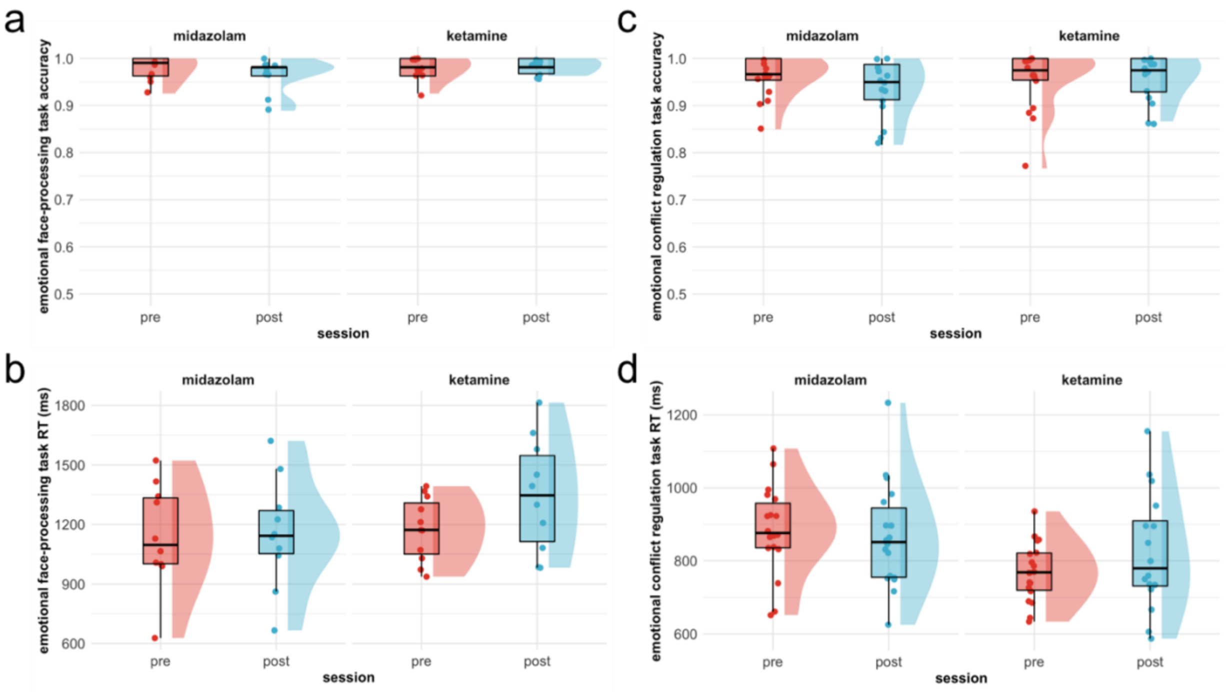


**Figure S1. Summary of behavioural performance on emotion-processing tasks, by imaging session (pre- *vs* post-infusions), and received drug (midazolam *vs* ketamine). a** Mean response accuracy on the emotional face-processing task. **b** Mean response times (RTs) on the emotional face-processing task. There was no evidence of an effect of session (pre- *vs* post-infusions) or drug (ketamine *vs* midazolam) on either face-matching accuracy or average response time measures (all $p>0.2$, repeated-measures ANOVAs). **c** Mean response accuracy on the emotional conflict regulation task (collapsed across congruent and incongruent trial types as there was no evidence accuracy varied according to task condition). **d** Mean response times on the emotional conflict regulation task (collapsed across congruent and incongruent trial types as there was no evidence RT varied according to task condition). There was marginal evidence for an interaction between drug and session on response accuracy ($p=0.04$), which appeared to be driven by a tendency towards lower accuracy on the post-infusion session in participants who had received midazolam. There was no evidence of an effect of session or drug on response times (all $p>0.1$). Raincloud plots were generated using [58].

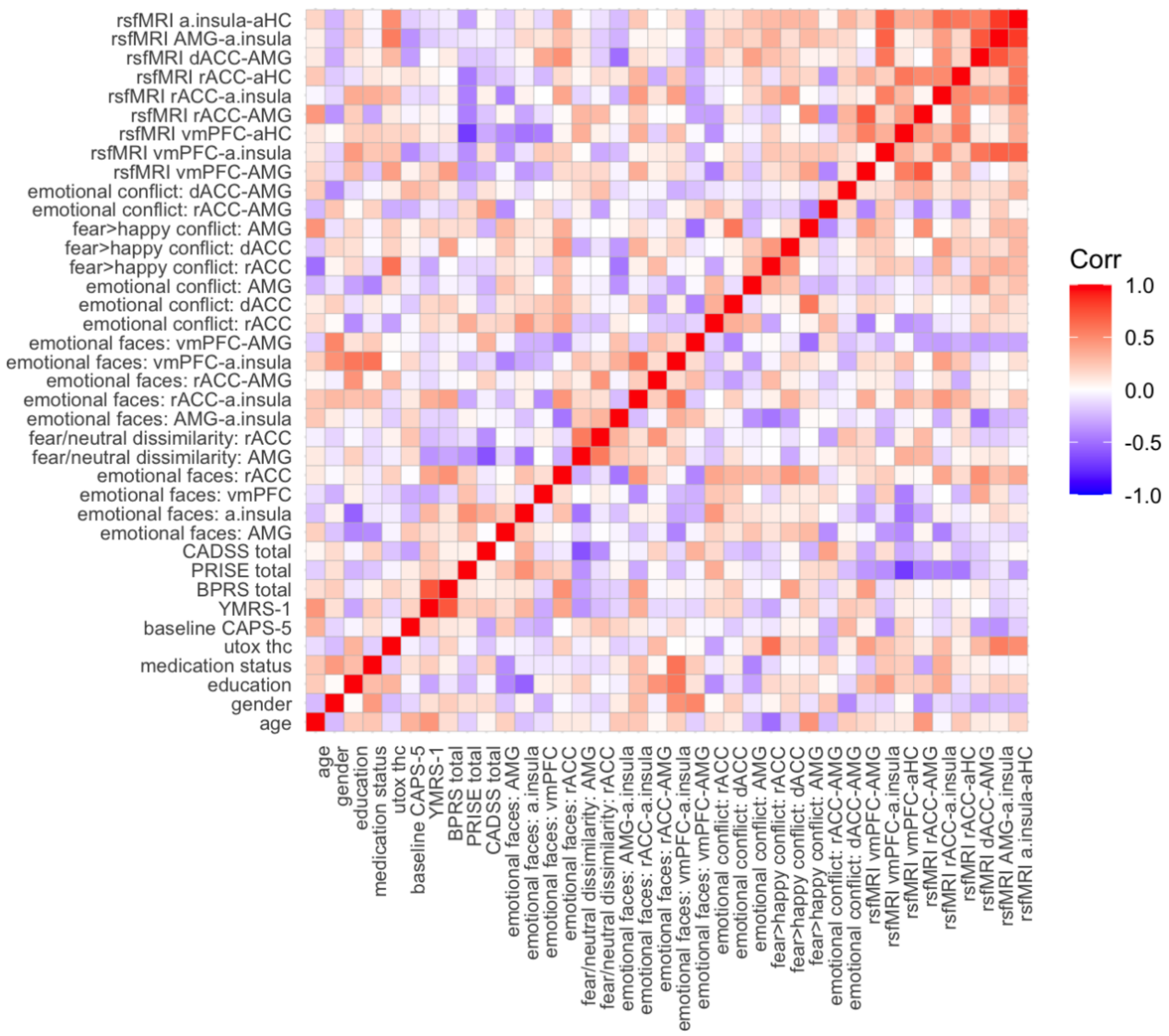


**Figure S2. Correlation matrix for variables included in the correlates of symptom change analysis**. Imaging measures represent change scores (post-infusion scan minus pre-infusion scan). rsfMRI, resting-state/task-free fMRI scan; a.insula, anterior insula; aHC, anterior hippocampus; AMG, amygdala; dACC, dorsal anterior cingulate cortex; rACC, rostral anterior cingulate cortex, vmPFC, ventromedial prefrontal cortex; CADSS, Clinician-Administered Dissociative States Scale; BPRS, four items from the Brief Psychiatric Rating Scale probing psychotomimetic symptoms; YMRS, a single item from the Young Mania Rating Scale indexing elevated mood; PRISE, Patient Rated Inventory of Side-Effects (total score calculated by summing across all somatic domains); CAPS-5, Clinician-Administered PTSD Scale for DSM-5; utox thc, urine toxicology results for presence of THC. Colour bar represents Pearson correlations.

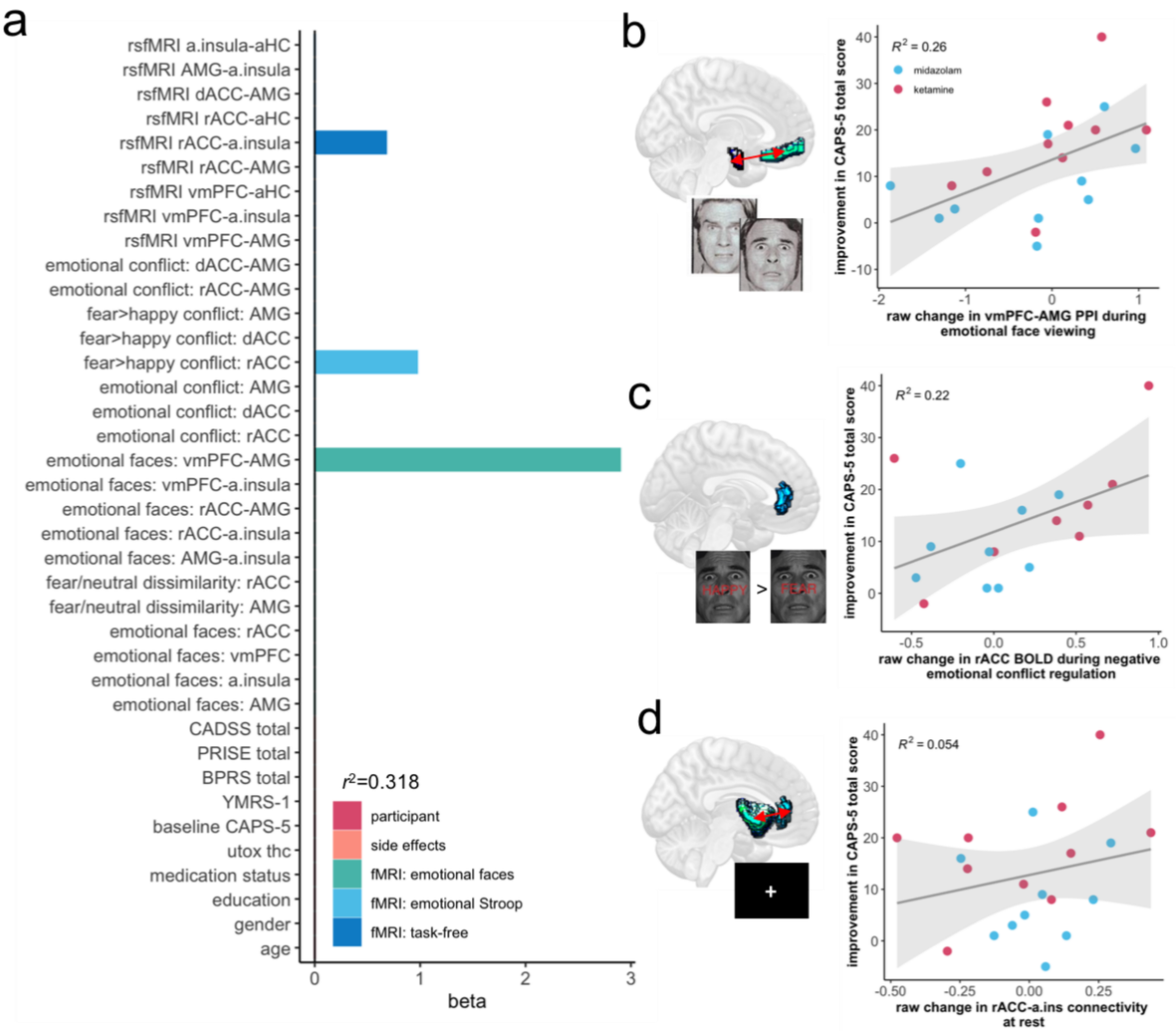


**Figure S3. Neuroimaging correlates of PTSD symptom change (drug-agnostic).** **a** Standardized regression coefficient (beta) values for the elastic net model with minimum predictive error for change in PTSD symptoms over the course of treatment in left-out subjects. Imaging measures represent change scores (post-infusion scan minus pre-infusion scan), across all participants (agnostic to received drug identity). rsfMRI, resting-state/task-free fMRI scan; a.insula, anterior insula; aHC, anterior hippocampus; AMG, amygdala; dACC, dorsal anterior cingulate cortex; rACC, rostral anterior cingulate cortex, vmPFC, ventromedial prefrontal cortex; CADSS, Clinician-Administered Dissociative States Scale; BPRS, four items from the Brief Psychiatric Rating Scale probing psychotomimetic symptoms; YMRS, a single item from the Young Mania Rating Scale indexing elevated mood; PRISE, Patient Rated Inventory of Side-Effects (total score calculated by summing across all somatic domains); CAPS-5, Clinician-Administered PTSD Scale for DSM-5; utox thc, urine toxicology results for presence of THC. **b** Increased connectivity between the vmPFC and amygdala during emotional face-viewing was the strongest predictor of improvement in total PTSD symptom severity. Increased in rACC BOLD during negative emotional conflict regulation (**c**) and increased resting rACC-a.insula functional connectivity (**d**) were also retained as predictors of PTSD symptom improvement in the minimum error model.

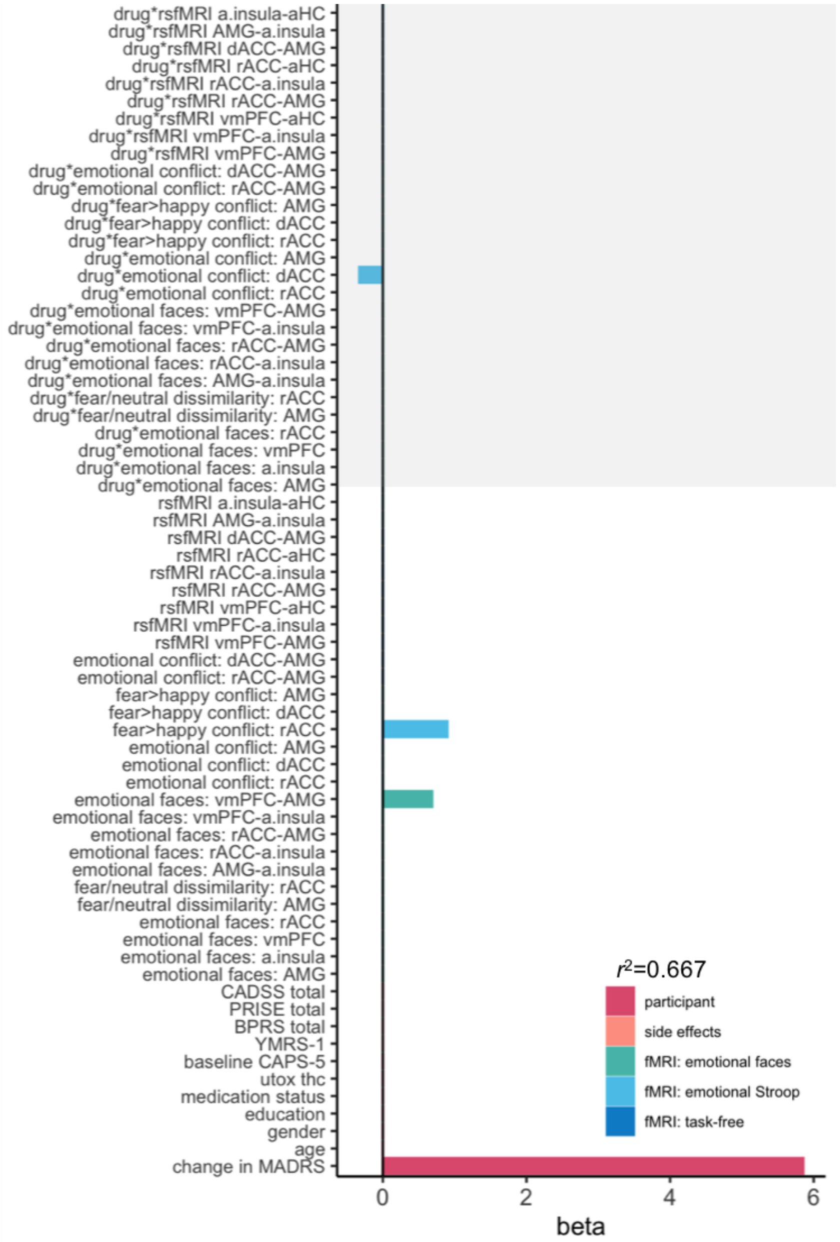


**Figure S4. Neuroimaging correlates of PTSD symptom change (including concomitant change in depression score).** **a** Standardized regression coefficient (beta) values for the elastic net model with minimum predictive error for change in PTSD symptoms over the course of treatment in left-out subjects. All imaging measures represent change scores (post-infusion scan minus pre-infusion scan). Grey shading highlights interaction terms between imaging measures and received drug identity (ketamine vs midazolam). rsfMRI, resting-state/task-free fMRI scan; a.insula, anterior insula; aHC, anterior hippocampus; AMG, amygdala; dACC, dorsal anterior cingulate cortex; rACC, rostral anterior cingulate cortex, vmPFC, ventromedial prefrontal cortex; CADSS, Clinician-Administered Dissociative States Scale; BPRS, four items from the Brief Psychiatric Rating Scale probing psychotomimetic symptoms; YMRS, a single item from the Young Mania Rating Scale indexing elevated mood; PRISE, Patient Rated Inventory of Side-Effects (total score calculated by summing across all somatic domains); CAPS-5, Clinician-Administered PTSD Scale for DSM-5; utox thc, urine toxicology results for presence of THC; MADRS, Montgomery-Åsberg Depression Rating Scale, total score.

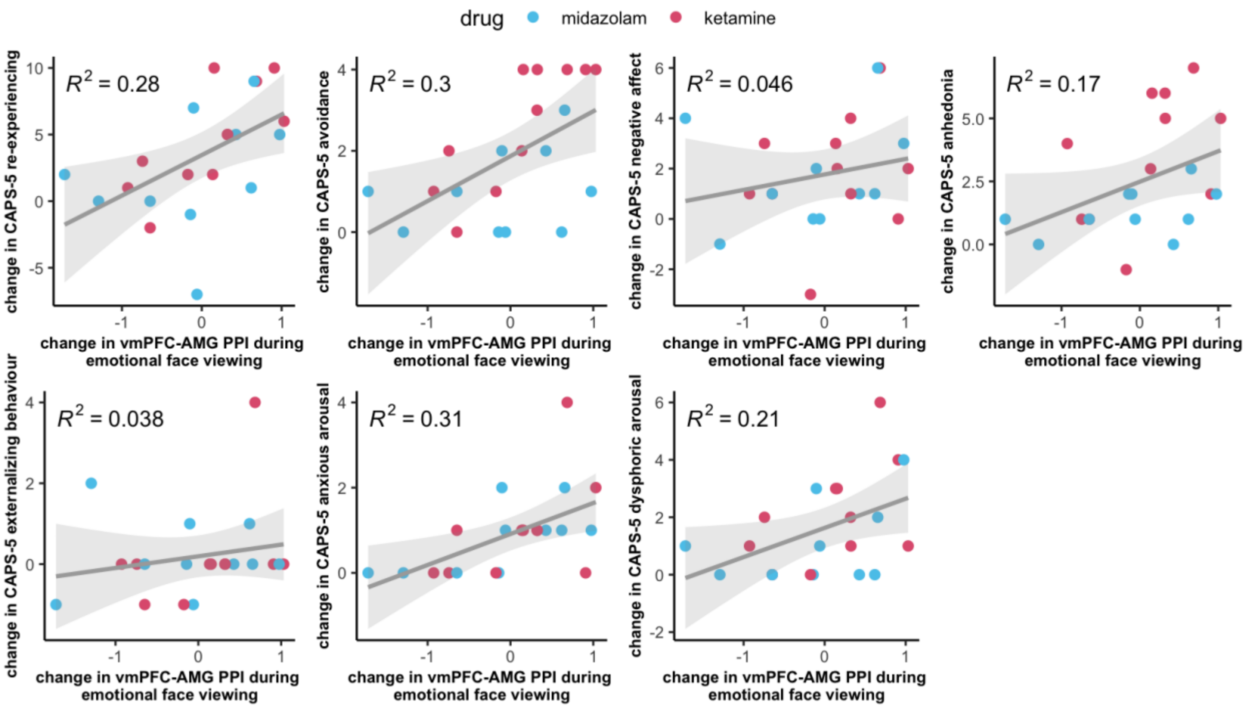


**Figure S5. Relationship between change in vmPFC-AMG functional connectivity during emotional face-viewing and change in PTSD symptoms, across PTSD symptom clusters**. Lines of best fit and *R2* values represent linear trends, with shading representing 95% CIs. PTSD symptom clusters are as defined by Armour et al. [59]. CAPS-5, Clinician-Administered PTSD Scale for DSM-5; vmPFC, ventromedial prefrontal cortex; AMG, amygdala; PPI, (generalized) psycho-physiological interaction analysis of task-specific functional connectivity between brain regions.

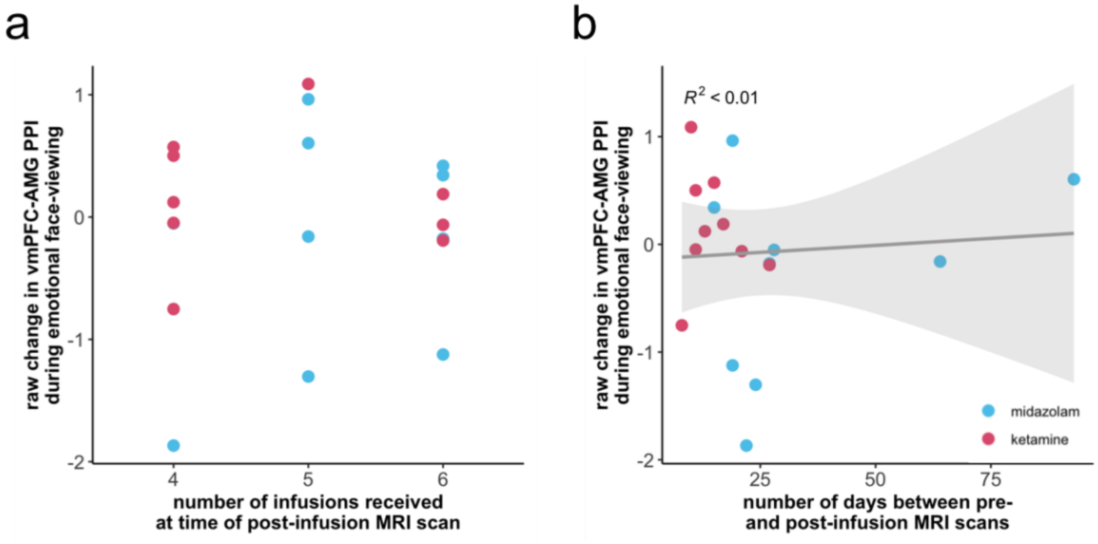


**Figure S6. Change in vmPFC-AMG functional connectivity during emotional face-viewing post- *vs* pre-infusion, according to number of infusions received at the time of the post-infusion MRI scan and time between scans in days. a** Due to scheduling conflicts, 2/3 participants had their second (post-infusion) MRI scan the day after their 4$^{th}$ or 5$^{th}$ infusion, and 1/3 participants within 48 hours of their 6$^{th}$ (and final) infusion. vmPFC, ventromedial prefrontal cortex; AMG, amygdala; PPI, (generalized) psycho-physiological interaction analysis of task-specific functional connectivity between brain regions. **b** The same data as in *a*, plotted by time (in days) between the baseline (pre-infusion) and post-infusion scan sessions. *NB*, for all participants, the post-infusion scan was carried out at the latest 48-hours following the 6$^{th}$ (final) infusion. The larger delays for some participants are due to the *pre*-infusion or baseline scan occurring some time before the initiation of the infusion regime (at which point in time, all participants were symptomatic for PTSD).

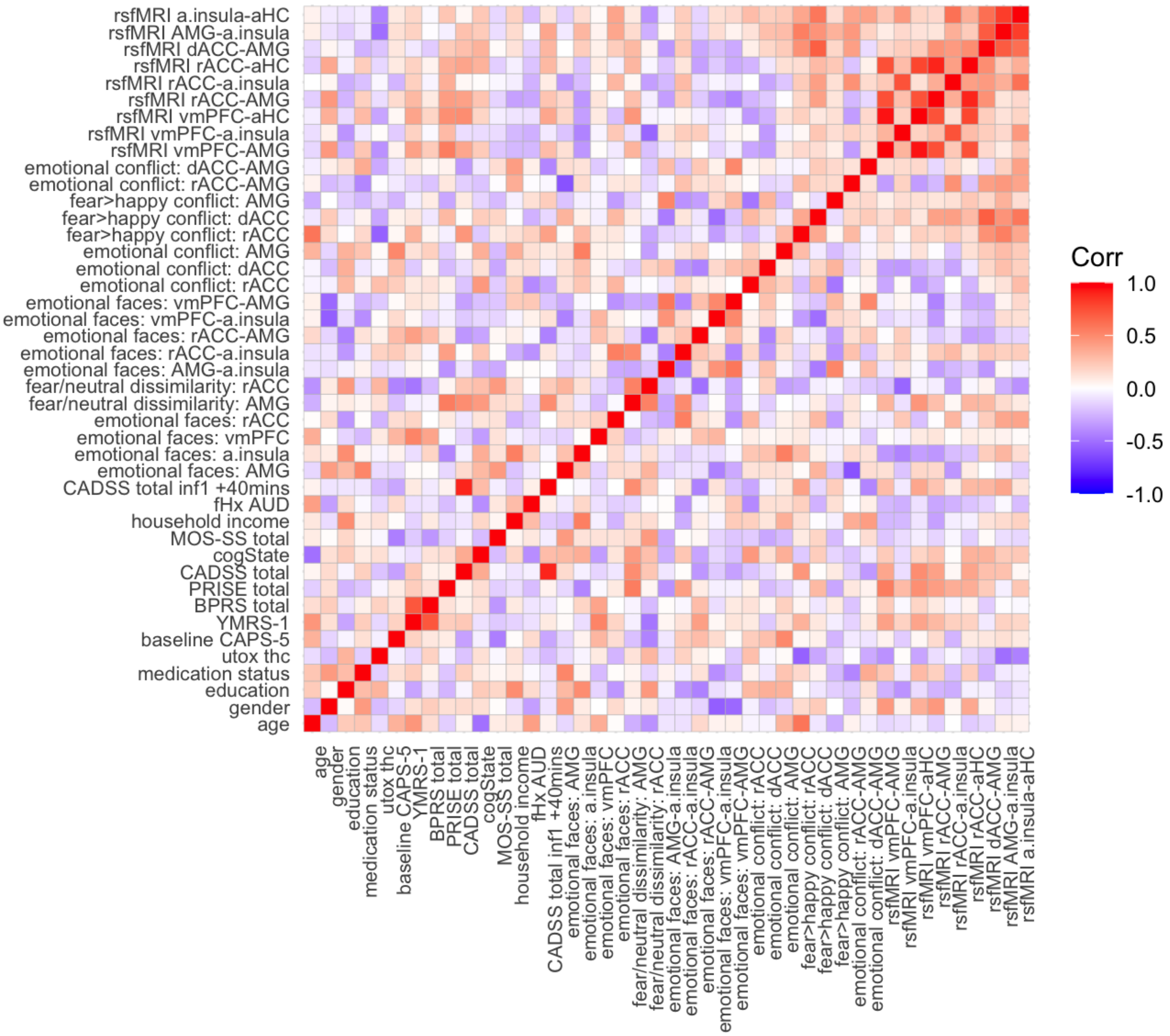


**Figure S7. Correlation matrix for variables included in the baseline prediction analysis**. Imaging measures represent estimates extracted from the baseline (pre-infusion) session. rsfMRI, resting-state/task-free fMRI scan; a.insula, anterior insula; aHC, anterior hippocampus; AMG, amygdala; dACC, dorsal anterior cingulate cortex; rACC, rostral anterior cingulate cortex, vmPFC, ventromedial prefrontal cortex; fHX AUD, family history of alcohol use disorders in first degree relatives; MOS-SS, Medical Outcomes Study Social Support Survey total score; cogState, composite score for executive function derived from the cogState neurocognitive test battery; CADSS, Clinician-Administered Dissociative States Scale; BPRS, four items from the Brief Psychiatric Rating Scale probing psychotomimetic symptoms; YMRS, a single item from the Young Mania Rating Scale indexing elevated mood; PRISE, Patient Rated Inventory of Side-Effects (total score calculated by summing across all somatic domains); CAPS-5, Clinician-Administered PTSD Scale for DSM-5; utox thc, urine toxicology results for presence of THC. Colour bar represents Pearson correlations.

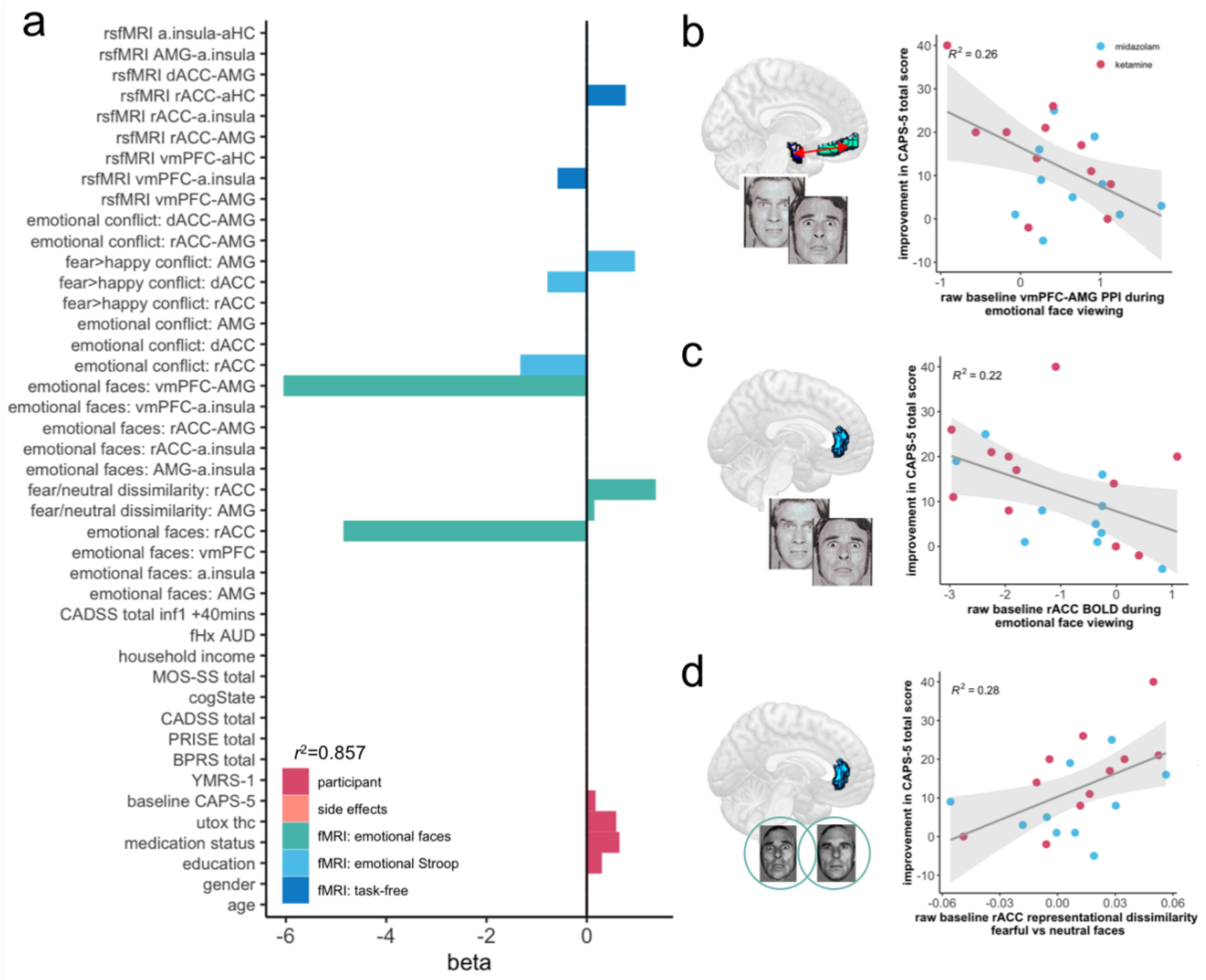


**Figure S8. Baseline predictors of PTSD symptom change (drug-agnostic). a** Standardized regression coefficient (beta) values for the elastic net model with minimum predictive error for change in PTSD symptoms over the course of treatment in left-out subjects. Imaging measures represent estimates extracted from the baseline (pre-infusion) session. rsfMRI, resting-state/task-free fMRI scan; a.insula, anterior insula; aHC, anterior hippocampus; AMG, amygdala; dACC, dorsal anterior cingulate cortex; rACC, rostral anterior cingulate cortex, vmPFC, ventromedial prefrontal cortex; fHX AUD, family history of alcohol use disorders in first degree relatives; MOS-SS, Medical Outcomes Study Social Support Survey total score; cogState, composite score for cognitive function derived from the cogState neurocognitive test battery; CADSS, Clinician-Administered Dissociative States Scale; BPRS, four items from the Brief Psychiatric Rating Scale probing psychotomimetic symptoms; YMRS, a single item from the Young Mania Rating Scale indexing elevated mood; PRISE, Patient Rated Inventory of Side-Effects (total score calculated by summing across all somatic domains); CAPS-5, Clinician-Administered PTSD Scale for DSM-5; utox thc, urine toxicology results for presence of THC. **b** Decreased baseline connectivity between the vmPFC and amygdala during emotional face-viewing was the strongest predictor of improvement in total PTSD symptom severity during treatment. Lower rACC BOLD during negative emotional face-viewing (**c**) and emotional conflict regulation, and increased baseline representational distance between fearful and neutral faces in the rACC (**d**) were also retained as baseline predictors of PTSD symptom improvement in the minimum error model.

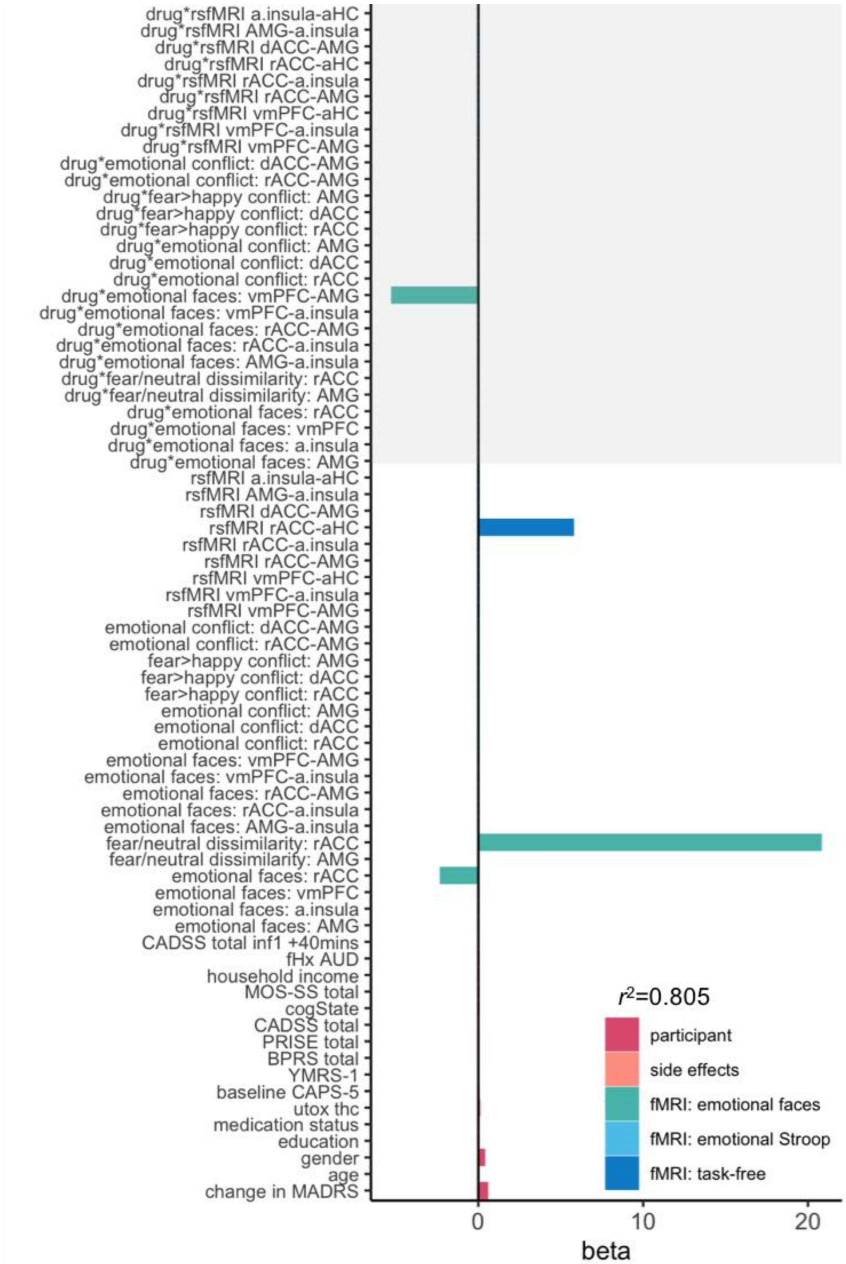


**Figure S9. Baseline predictors of PTSD symptom change (including concomitant change in depression score)**. **a** Standardized regression coefficient (beta) values for the elastic net model with minimum predictive error for change in PTSD symptoms over the course of treatment in left-out subjects. Imaging measures represent estimates extracted from the baseline (pre-infusion) session. Grey shading highlights interaction terms between imaging measures and received drug identity (ketamine vs midazolam). rsfMRI, resting-state/task-free fMRI scan; a.insula, anterior insula; aHC, anterior hippocampus; AMG, amygdala; dACC, dorsal anterior cingulate cortex; rACC, rostral anterior cingulate cortex, vmPFC, ventromedial prefrontal cortex; fHX AUD, family history of alcohol use disorders in first degree relatives; MOS-SS, Medical Outcomes Study Social Support Survey total score; cogState, composite score for executive function derived from the cogState neurocognitive test battery; CADSS, Clinician-Administered Dissociative States Scale; BPRS, four items from the Brief Psychiatric Rating Scale probing psychotomimetic symptoms; YMRS, a single item from the Young Mania Rating Scale indexing elevated mood; PRISE, Patient Rated Inventory of Side-Effects (total score calculated by summing across all somatic domains); CAPS-5, Clinician-Administered PTSD Scale for DSM-5; utox thc, urine toxicology results for presence of THC.

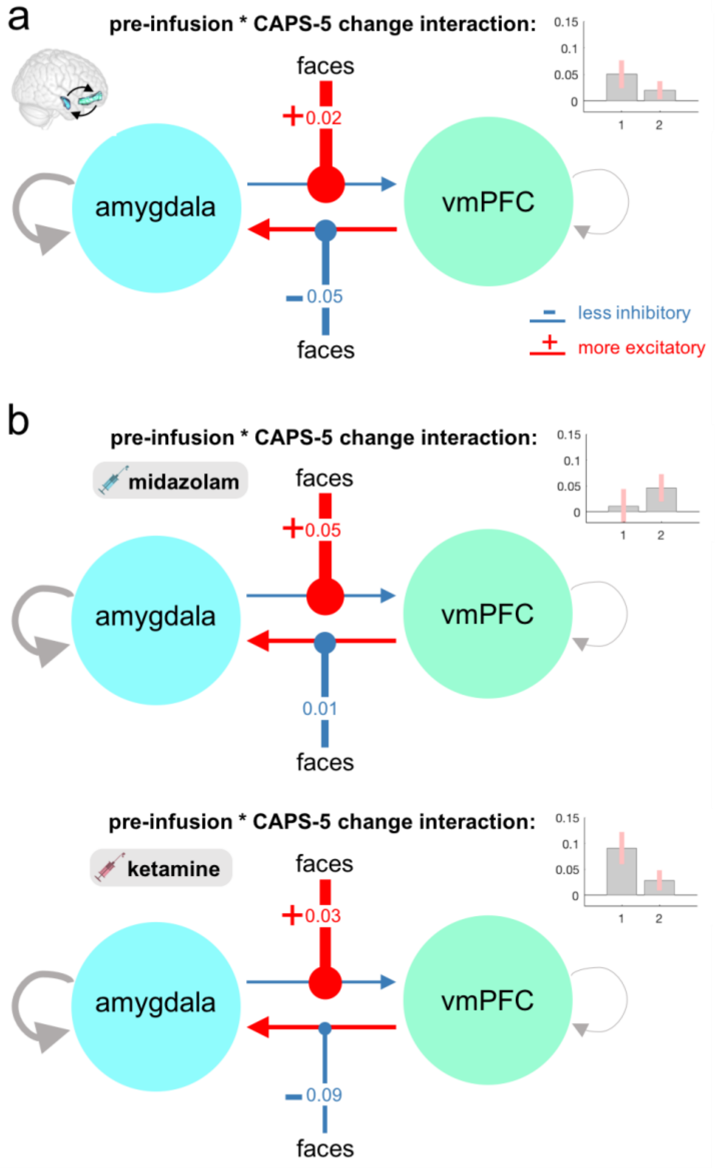


**Figure S10. Dynamic causal modelling of task-modulated effective connectivity during the emotional face-viewing task (baseline prediction analysis)**. **a** Parametric Empirical Bayes (PEB) analysis of pre-infusion effective connectivity related to response to treatment (future improvement in CAPS-5 total scores) revealed that across all participants, improvement in overall PTSD severity was associated with decreased baseline vmPFC inhibition of the amygdala, and increased amygdala excitation of the vmPFC, during emotional face viewing. **b** When divided by drug, the relationship between future PTSD symptom improvement and decreased baseline top-down inhibition of the amygdala by the vmPFC during emotional stimuli was only evident in participants who went on to receive ketamine, a difference confirmed by passing both models to a 3rd level PEB analysis (more positive parameter estimate for the vmPFC→amygdala connection in the ketamine group; posterior probability=0.95). For all panels, pointed arrowheads represent connections between regions, and circular arrowheads represent modulation of those connections by the experimental condition of interest (emotional faces). Values are rates of change constants in Hz. Insets depict PEB posterior parameter estimates for modulation of connectivity by the listed effect of interest: 1, vmPFC→amygdala; 2, amygdala→vmPFC; error bars represent 90% confidence intervals.
